# Estimating disease heritability from complex pedigrees allowing for ascertainment and covariates

**DOI:** 10.1101/2023.07.13.23292588

**Authors:** Doug Speed, David M. Evans

## Abstract

We propose TetraHer, a method for estimating the liability heritability of binary phenotypes. TetraHer has five key features. Firstly, it can be applied to data from complex pedigrees, that contain multiple types of relationships. Secondly, it can correct for ascertainment of cases or controls. Thirdly, it can accommodate covariates. Fourthly, it can model the contribution of common environment. Fifthly, it produces a likelihood, that can be used for significance testing. We first demonstrate the validity of TetraHer on simulated data. We then use TetraHer to estimate liability heritability for 229 codes from the tenth International Classification of Diseases (ICD-10). We identify 118 codes with significant heritability (P<0.05/229), which can be used in future analyses for investigating the genetic architecture of human diseases.

## INTRODUCTION

Estimates of heritability are of great value in statistical genetics. For example, they indicate the value of performing a genetic association study, and provide an upper bound for the accuracy of genetic prediction models.^1,2^ For a quantitative trait, heritability is defined as the proportion of phenotypic variation explained by genetic factors.^3^ For a binary phenotype, the same definition is referred to as heritability on the observed scale. An alternative is to assume a liability model, and then consider the proportion of liability variation explained by genetic factors, referred to as heritability on the liability scale.^4^ Most methods for estimating heritability were originally designed for quantitative traits. However, these methods are often applied to binary phenotypes via a two-step approach; first the method is used to estimate heritability on the observed scale, then this estimate is converted to the liability scale via a linear transformation.^5,6^

In this paper, we focus on binary disease phenotypes, where each individual is recorded as either affected (a case) or unaffected (a control). When analyzing diseases, we generally prefer heritability estimates on the liability scale, because these do not depend on the prevalence and ascertainment of the disease, and can be readily compared across studies and traits.^4,7^

We begin by using simulations to evaluate four existing methods for estimating liability heritability. The first two methods, Pearson’s correlation and REML, use a two-step approach and tend to produce upwardly-biased estimates.^3,8^ The bias is highest for diseases with substantial heritability and low prevalence. For example, for diseases with prevalence 1%, the estimate can be more than twice the true value. We show that the bias arises because the linear transformation from the observed to liability scale fails for close relatives. The third method, PCGC, estimates liability heritability directly, but nonetheless exhibits biases similar to those of the two-step approaches.^9^ By contrast, the fourth method, tetrachoric correlation, estimates liability heritability directly and produces unbiased estimates.^10^

Our simulation results motivate us to develop TetraHer, which addresses five limitations of tetrachoric correlation. Firstly, TetraHer can be applied to complex pedigrees, where there are a mix of different relationships (including “non-standard” relationships inferred from SNP data). Secondly, TetraHer can correct for ascertainment, and thus produce more accurate estimates of liability heritability when cases or controls have been over-sampled. Thirdly, TetraHer can accommodate covariates. Fourthly, TetraHer can model the contribution of common environment. Fifthly, TetraHer reports a likelihood, which can be used to test whether the estimated heritability (or contribution of common environment) is significant.

We use TetraHer to estimate the heritability of 229 ICD-10 codes with prevalence at least 2% in the UK Biobank.^11,12^ We find that 118 of the codes have significant heritability (P<0.05/229), spanning 12 disease chapters. We then use these 118 codes to investigate the relationship between per-SNP heritability and minor allele frequency (MAF).^13–15^

## METHODS

Here we summarize the methods and data used in this paper, with full details provided in **Supplementary Notes 1, 2 & 3**. Be aware that in the mathematical details, we use square brackets to specify elements of a vector or matrix (e.g., a_1_[b] denotes the bth element of a vector called a_1_).

### Notation

Suppose we have a sample of n individuals, and let the length-n vector Y record which are affected (Y[i]=1) and unaffected (Y[i]=0) for a particular disease. Let A=mean(Y[i]) denote the ascertainment of the disease (the proportion of cases in the sample), and let K denote the prevalence of the disease (the proportion of cases in the population). Further, suppose we have p covariates, whose values are contained within the n x p matrix Z. Throughout this paper, we assume a liability threshold model, as described in **Figure 1a**. If the length- n vector L denotes the (unobserved) liabilities of individuals, then Y[i]= I(L[i]>T), where T=Φ^-1^(K) is the (1-K)th quantile of a standard normal distribution (e.g., if K=0.01, then Y[i] indicates whether or not L[i] is greater than 2.32).

**Figure 1:**
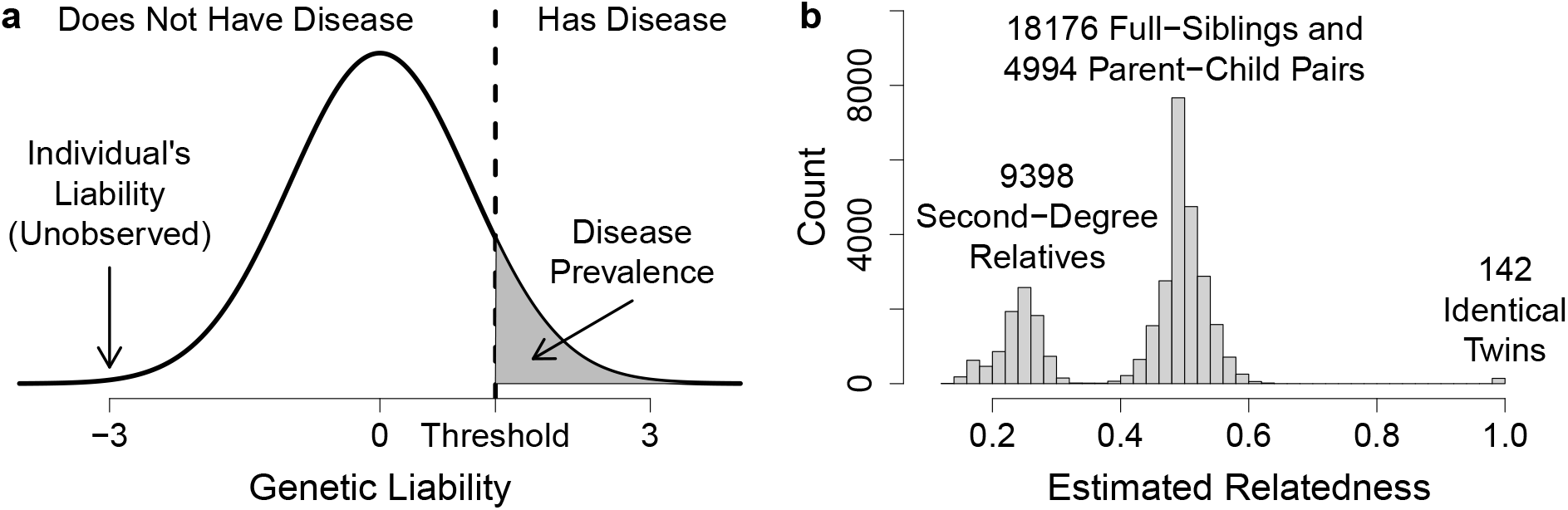
Liability model and distribution of related pairs in the UK Biobank. (a) The liability model assumes that an individual’s disease status indicates whether their liability (an unobserved, standard normally distributed random variable) is above (affected) or below (unaffected) a threshold (determined by the prevalence of the disease). (b) The distribution of SNP-derived estimates of relatedness for the 32,710 white British, related pairs in the UK Biobank.

Let R denote a symmetric n x n matrix such that R[i,j] records the genetic similarity between Individuals i and j. Traditionally, R contained coefficients of relatedness, the expected proportions of genome sharing, derived from pedigree information (e.g., full-siblings have R[i,j]=0.5, half-siblings have R[i,j]=0.25, etc). However, it is now common for R to contain SNP-derived estimates of relatedness, which measure the actual proportion of genome sharing (e.g., while full-siblings are expected to share half their genome, **Figure 1b** shows that the actual proportion shared typically ranges from 0.4 to 0.6).^16–18^

### Heritability definitions

When modeling variation in the observed phenotypes, we assume Y = F + G + C + E, where the independent, length-n vectors F, G, C and E denote the contributions of covariates, genetic factors, common environment and environmental noise, respectively. We then define heritability on the observed scale as h^2^_O_ =Var(G)/(Var(Y)-Var(F)). When modeling variation in the liabilities, we instead assume L= F + G + C + E, and define heritability on the liability scale as h^2^_L_ =Var(G)/(Var(L)-Var(F)). Most existing heritability methods are designed to estimate h^2^_O_. However, it has been argued that these estimates can be converted to the liability scale via the transformation

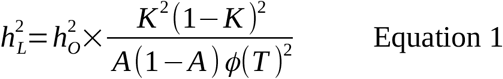

where φ(T) is the density of a standard normal distribution evaluated at the threshold.^4,6^

Note that the above definitions of h^2^_O_ and h^2^_L_ have been referred to as “conditional”, because the denominators denote the variance of the phenotype or liability after allowing for covariates.^9^ An alternative is to consider the “marginal” heritabilities Var(G)/Var(Y) and Var(G)/Var(L). We prefer conditional heritabilities, because in our opinion, heritability is ideally defined with respect to a homogeneous population, and therefore we consider the covariates nuisance parameters. However, for completeness, we also compute and report marginal heritabilities.

Analogous to the above definitions of heritability, we let h^2^_C_ =Var(C)/(Var(L)-Var(F)) denote the proportion of liability variation explained by common environment.

### Existing methods

We consider four existing methods for estimating liability heritability: Pearson’s correlation and REML are two-step approaches (i.e., first obtain an estimate of h^2^_O_, then use Equation 1 to convert this to an estimate of h^2^_L_), while PCGC and Tetrachoric correlation estimate h^2^_L_ directly.^3,8–10^ Here we briefly describe the four methods, and for simplicity, we ignore covariates (fuller descriptions are provided in **Supplementary Note 2**). Pearson’s correlation estimates h^2^_O_ based on ρ_P_, the Pearson’s correlation between phenotypes of related pairs (e.g., its estimate of h^2^_O_ from full-siblings would be 2ρ_P_). REML estimates h^2^_O_ by assuming Y ∼N(0, RVar(G) + IVar(E)), where I is an n x n identity matrix, then finds Var(G) and Var(E) that maximize the (restricted) likelihood. PCGC constructs Y’, a standardized version of Y such that E(Y’[i]Y’[j]) ≈ R[i,j] h^2^_L_, then estimates h^2^_L_ by regressing the observed values of Y’[i]Y’[j] on R[i,j]. Tetrachoric correlation estimates h^2^_L_ directly based on ρ_T_, the tetrachoric correlation between phenotypes of related pairs (e.g., its estimate of h^2^_L_ from full-siblings would be 2ρ_T_).

### TetraHer

We first describe TetraHer assuming that the sample is not ascertained (i.e., A=K), and that there are no contributions from either covariates or common environment (i.e., F=0 and C=0); we then relax each of these conditions in turn. Suppose there are D related pairs (e.g., pairs with R[i,j]>0.05), and let the length-D vectors S_1_ and S_2_ index the first and second individuals in each pair, respectively. So for example, if Individuals 1 & 2 are related, we could set S_1_[i]=1 and S_2_[i]=2 (note that the order is arbitrary, so it is equivalent to instead set S_1_[i]=2 and S_2_[i]=1). In the following explanation, we use the vectors y_1_ and y_2_ to denote the phenotypes of the first and second individuals in each pair (i.e., y_1_=Y[S_1_] and y2=Y[S_2_]), use the vectors l_1_ and l_2_ to denote their liabilities (i.e., l1=L[S_1_] and y_2_=y[S_2_]), and use the vector r to denote their relatedness estimates (i.e., r[d] is the estimated relatedness between the dth pair).

TetraHer estimates h^2^_L_ by finding the value that maximizes the likelihood of the pairs of observed phenotypes. To construct a likelihood for the phenotype pair (y_1_[d], y_2_[d]), TetraHer assumes that the corresponding liability pair (l_1_[d], l_2_[d]) is a draw from a bivariate standard normal distribution with correlation v[d]=r[d] h^2^_L_. It is then possible to compute P_00_[d], P_10_[d], P_01_[d] and P_11_[d], the probabilities of observing the phenotype pairs (0,0), (1,0), (0,1) and (1,1), respectively. For example,

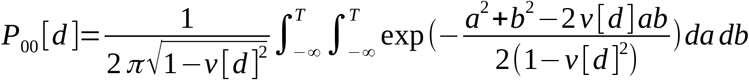

TetraHer computes a joint log likelihood by assuming the D pairs are independent

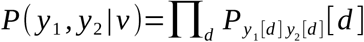

then estimates h^2^_L_ by maximizing the log likelihood using the Newton-Raphson Method. It estimates the variance of the estimate by inverting the second derivative of the log likelihood.

When the sample is ascertained (i.e., A≠K), TetraHer revises how it computes the probabilities of observing the four different phenotype pairs. For example, it now computes

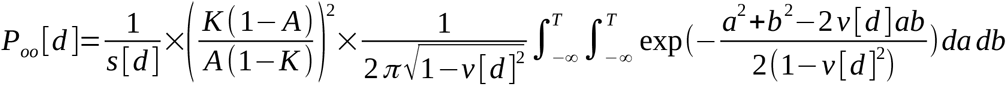

where the scalar s[d] ensures that P_00_[d], P_10_[d], P_01_[d] and P_11_[d] sum to one. The ratio (K(1-A))/(A(1-K)) is the relative probability that an unaffected individual is included in the sample (it will be less than one if A>K, and vice versa).

When allowing for covariates (i.e., F≠0), TetraHer starts by obtaining F’, an estimate of F. Copying the approach of PCGC, TetraHer uses logistic regression to estimate the probabilities that individuals are affected given their covariates, then converts these to an estimate of F in a way that allows for ascertainment. ^9^ If f_1_ and f_2_ contain the covariates estimates for the first and second individuals in each pair (i.e., f_1_=F’[S_1_] and f_2_=F’[S_2_]), then TetraHer assumes that l_1_[d] - f_1_[d] and l_2_[d] - f_2_[d] are draws from a bivariate standard normal distribution with correlation r_d_ h^2^_L_ (i.e., replaces liabilities with their values after adjusting for covariates).

When allowing for common environment (i.e., C≠0), TetraHer assumes (l_1_[d]- f_1_[d], l_2_[d]- f_2_[d]) is a draw from a bivariate standard normal distribution with correlation v[d] = r[d] h^2^_L_ + c[d] h^2^_C_, where the length-D vector c describes the degree of common environment for the related pairs. Note that it is only possible to obtain (sensible) estimates of h^2^_C_ when r and c are not linearly dependent.

In addition to TetraHer, we have also developed QuantHer, which is the analogous method for continuous phenotypes (e.g., when assuming no contributions from either covariates or common environment, QuantHer constructs a likelihood by assuming the phenotype pair (y_1_[d], y_2_[d]) is a draw from a bivariate normal distribution, with correlation r[d] h^2^_O_).

### Relationship between TetraHer and tetrachoric correlation

We consider TetraHer a generalization of tetrachoric correlation. Specifically, **Supplementary Figure 1** shows that if we apply TetraHer to pairs of individuals with the same relationship (i.e., where r_d_=r_1_), and assume no ascertainment, nor contributions from either covariates or common environment (i.e., assume A=K, F=0 and C=0), then the resulting estimates of h^2^_L_ almost exactly equal ρ_T_/r_1_ (where we obtain ρ_T_ using the R package polycor^19,20^).

### Similarities between TetraHer and PCGC

TetraHer assumes the same model as PCGC, however, the two methods differ in their solvers.^9^ Instead of computing the probabilities P_00_[d], P_10_[d], P_01_[d] and P_11_[d] exactly, PCGC uses an approximation that relies on v[d] being small. PCGC is designed for analyzing unrelated pairs (e.g., r_d_<0.05), in which case v[d] will tend to be very small, and the approximation is reasonable. However, when applied to related pairs, v[d] will often be substantial, and this approximation performs poorly (leading to the the biases observed below).

### Similarities between TetraHer and structural equation modelling

In addition to the four existing methods described above, it is also possible to estimate liability heritability via structural equation modeling (SEM).^21^ In **Supplementary Figure 2**, we show that for the simplest analysis (i.e., when r_d_=r_1_, and assuming A=K, F=0 and C=0) estimates of h^2^_L_ from SEM are almost identical to those from TetraHer. We believe that, in theory, many of the features of TetraHer are possible within SEM. However, we found that, despite trying alternative SEM software (e.g., lavaan,^22^ OpenMx^23^ and sem^24^), it was challenging to incorporate many of the features of TetraHer (e.g., allowing for complex pedigrees or ascertainment). We provide further comparison of TetraHer and SEM in the Discussion.

### Data

In total, the UK Biobank contains approximately 487k individuals.^11,12^ We first restrict to the 397,987 individuals who self-identified as white British, and whom we inferred to have European ancestry (via principal component analysis). We then used the software KING to infer family relationships.^25^ This identified 32,710 pairs of individuals within two degrees (142 identical twins, 18,176 full-siblings, 4,994 parent-child pairs, and 9,398 second-degree relatives), that span 56,602 unique individuals (**Figure 1b**). Each of these individuals is recorded for 23 covariates: age, sex, Townsend Deprivation Index and 20 principal components.

We first use the UK Biobank data for simulations (i.e., to generate phenotypes where we know the true heritability), then to estimate the heritability of diseases defined by ICD-10 codes (field 41270). In total, there are 19,133 ICD-10 codes, which are divided into 22 chapters (i.e., types of disease) and four levels (e.g., Level 3 codes are sub-categories of Level 2 codes). We restrict to the 229 codes in Chapters 1-15 with prevalence in the UK Biobank of at least 2%, of which 65, 90 and 74 are in Levels 1, 2 and 3, respectively. Note that for 37 of the codes, at least 80% of affected individuals were the same sex (females were predominantly affected for 29 codes, while males were predominantly affected for 8 codes), so for these we exclude individuals of the less-common sex in all analyses.

When running REML, PCGC and TetraHer, we set R based on the kinship estimates from KING (REML and PCGC require R_ij_ for all pairs of individuals, so we set R_ij_=0 for pairs that King does not infer to be related). Pearson’s correlation and Tetrachoric correlation can only be applied to pairs of individuals with the same relatedness. Therefore, we run each method twice for each phenotype, first using 23,170 pairs of full-siblings and parent-children (R_ij_=0.5), then using 9,398 second-degree relatives (R_ij_=0.25). We then combine the two estimates of h^2^_L_ into a single estimate via inverse-variance weighting.

### Software

We make TetraHer available within our software package LDAK.^18^ Note that TetraHer is computationally efficient. For example, in the analyses below, we have approximately 30,000 pairs of related individuals, and TetraHer completes in seconds, and this remains the case even with 100,000s of related pairs.

Furthermore, we have designed TetraHer so it is easy to use. In particular, all analyses in this paper can be performed using a one-line command, and we have ensured that, when possible, the TetraHer syntax matches that used by the popular software PLINK^26^ (for example, phenotypes and covariates are specified using the flags–pheno and –covar, respectively). We provide full instructions for running TetraHer (including test datasets) on the LDAK website, with a summary in **Supplementary Note 4**.

## RESULTS

### Advantage of tetrachoric correlation and TetraHer over two-step methods

First we simulate diseases with prevalence 1%, 10% or 50%, with h^2^_L_ equal to 0.2, 0.5 or 0.8, with no ascertainment, nor contributions from either covariates or common environment. **Figure 2a, 2b & 2c** show that estimates of h^2^_L_ from Pearson’s correlation, REML and PCGC are upwardly biased, with the extent of the bias depending on the heritability and prevalence. For example, for traits with prevalence 1% and heritability 0.5 or 0.8, the average estimate of h^2^_L_ is over twice the true value. By contrast, tetrachoric correlation and TetraHer produce consistent estimates of h^2^_L_.

**Figure 2:**
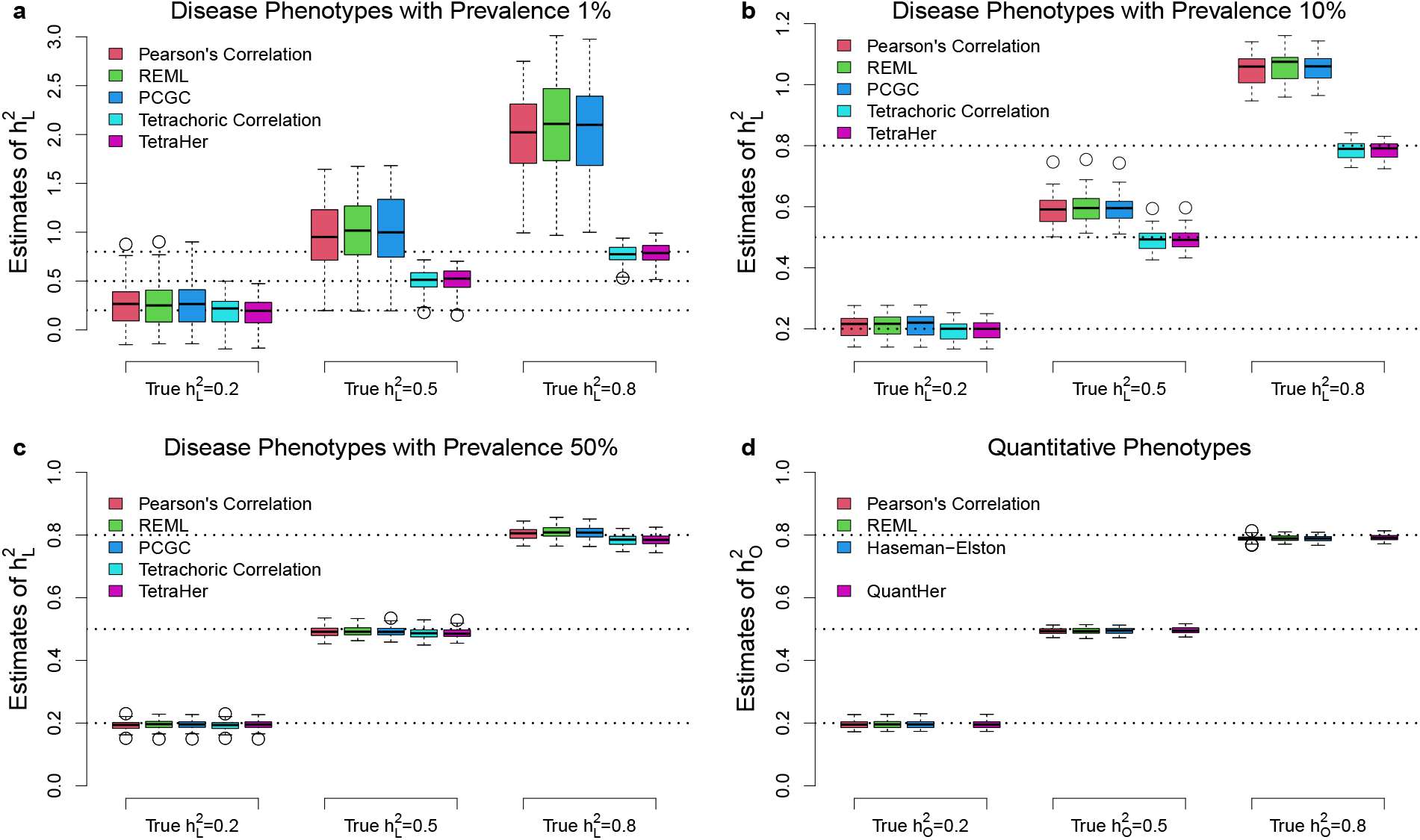
Comparison of methods for estimating heritability of simulated phenotypes. For (a), (b) and (c), we simulate disease phenotypes with prevalence 1%, 10% and 50%, respectively, then estimate liability heritability, h^2^_L_, using Pearson’s correlation, REML, PCGC, Tetrachoric correlation and TetraHer; for (d), we simulate quantitative phenotypes, then estimate observed heritability, h^2^_O_, using Pearson’s correlation, REML, Haseman-Elston regression and QuantHer. Boxes report estimates across 50 replicates (horizontal lines mark the 25^th^, 50^th^ and 75^th^ percentiles). Dashed horizontal lines indicate the true heritability (0.2, 0.5 or 0.8, depending on phenotype).

For the above simulations, the diseases are moderately polygenic (1000 causal SNPs), and effect sizes are sampled such that causal SNPs with lower MAF tend to explain less phenotypic variation (a tendency observed for real human traits).^27^ However, **Supplementary Figures 3 & 4** show that the results are almost identical if we instead consider highly polygenic diseases (20,000 causal SNPs) or generate effect sizes so that all causal SNPs are expected to explain equal phenotypic variation.

**Figure 2d** show that when applied to simulated quantitative traits, Pearson’s correlation, REML and Haseman-Elston regression^28^ (the equivalent of PCGC for quantitative traits) produce unbiased estimates of h^2^_0_. This indicates that the inflation observed for binary phenotypes occurs when converting heritability estimates from the observed to liability scale.^10^ **Figure 3** shows that the inflation arises because Equation 1 is only a good approximation when the correlations between pairwise liabilities (equal to r[d] h^2^_L_ +c[d] h^2^_C_) tend to be small, or when the prevalence of the disease is close to 50%.

**Figure 3.**
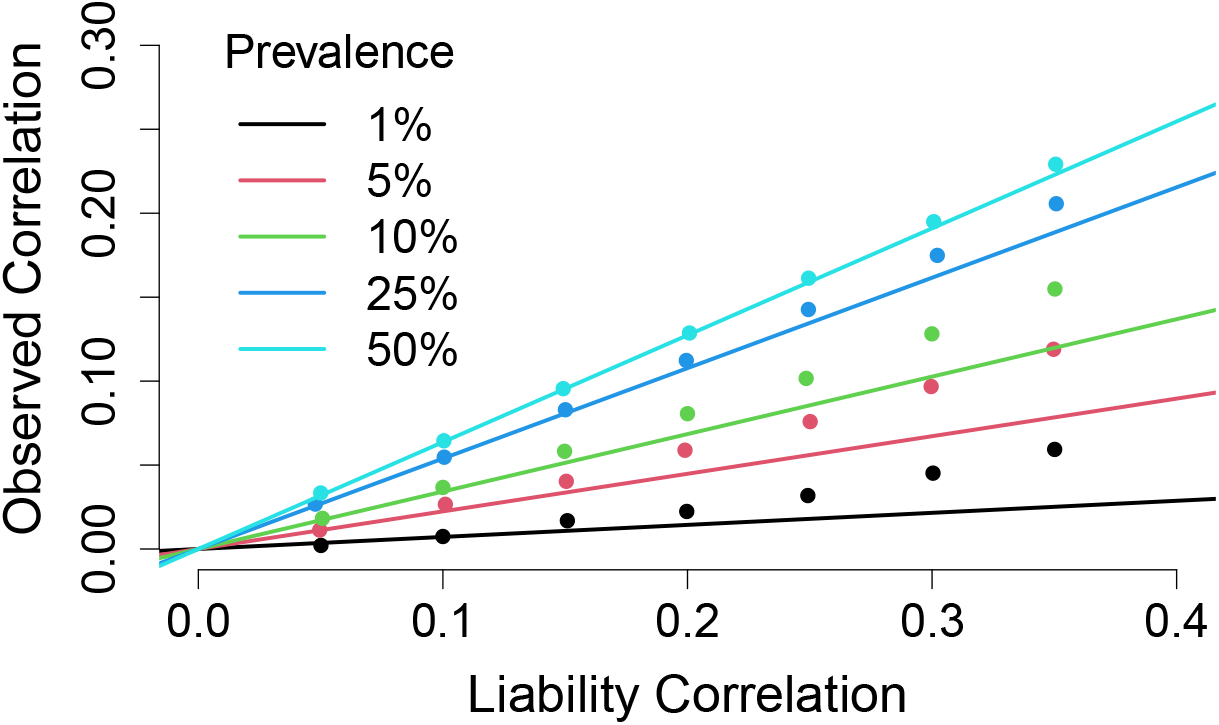
Relationship between correlations on the observed and liability scales. We generate pairs of liabilities with correlations ranging from 0.05 to 0.4, then convert these to pairs of binary phenotypes with prevalences between 1% and 50%. The points show how the correlation between the binary phenotypes (y-axis) depends on the correlation between the corresponding liabilities (x-axis) and the prevalence, while the lines show the relationship predicted by Equation 1.

### Advantages of TetraHer over tetrachoric correlation

TetraHer is able to analyze all pairs of individuals together, whereas tetrachoric correlation can only analyze pairs with the same relatedness. This is primarily a convenience (i.e., TetraHer can analyze all data in a single analysis, instead of multiple, and it can accommodate non-standard estimates of relatedness). However this feature also leads to a slight increase in precision. For example, **Figure 4a** shows that for diseases with h^2^_L_=0.5 and prevalence 1%, estimates from TetraHer have standard deviations on average 10% smaller than those from tetrachoric correlation.

**Figure 4:**
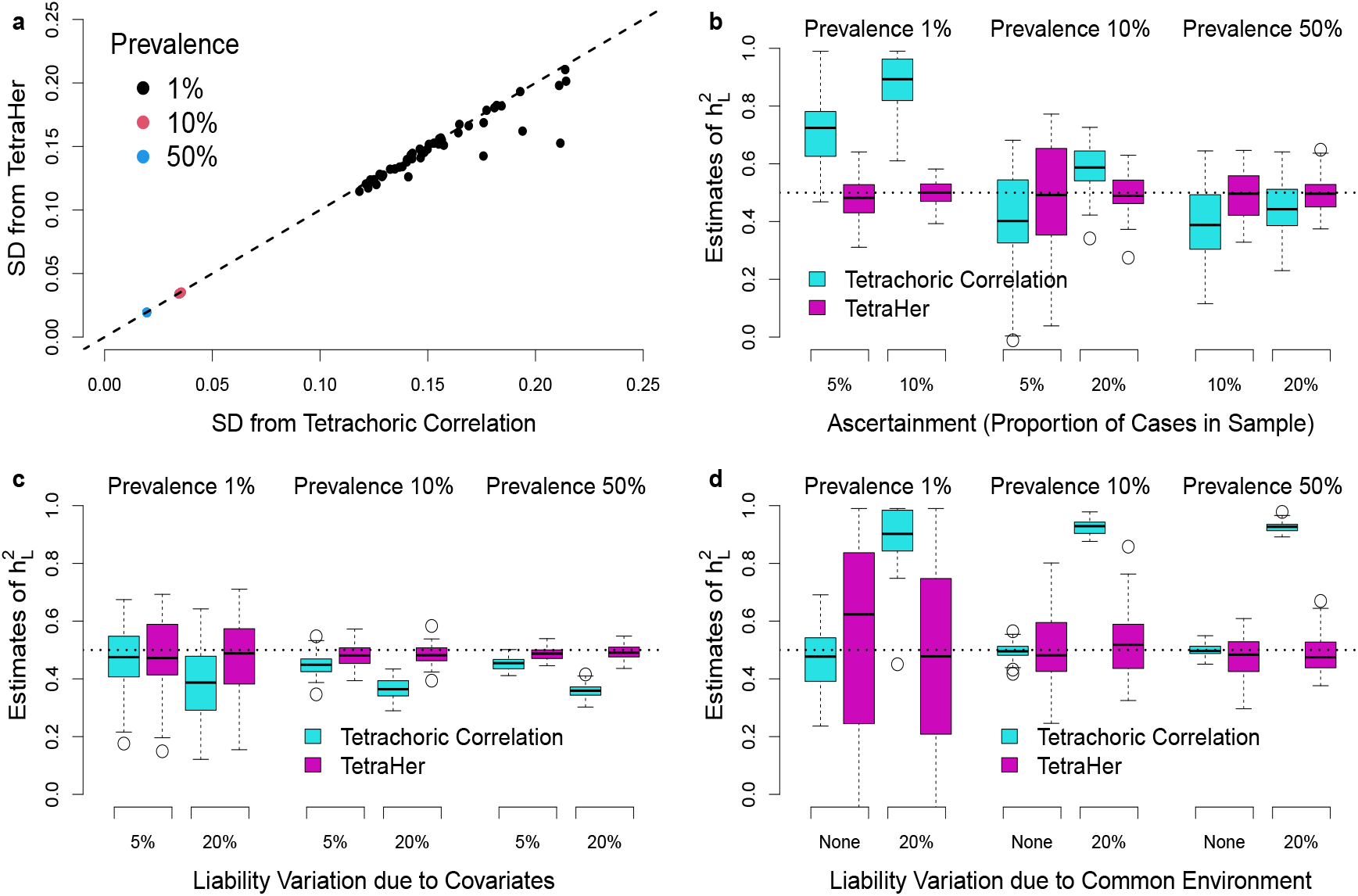
Comparison of tetrachoric correlation and TetraHer on simulated diseases. (a) We first simulate disease phenotypes with h^2^_L_ =0.5 and prevalence 1%, 10% or 50%. Points compare the standard deviation of estimates of h^2^_L_ from tetrachoric correlation and TetraHer. We then modify the phenotypes in three ways: (b) so that the proportion of cases in the sample no longer matches the disease prevalence, (c) so that the covariate age explains either 5% of 20% of liability variation, (d) so that common environment explains either 0% or 20% of liability variation. Boxes report estimates of h^2^_L_ across 50 replicates (horizontal lines mark the 25^th^, 50^th^ and 75^th^ percentiles).

For **Figure 4b**, we introduce ascertainment, by either over-sampling or under-sampling cases. TetraHer can allow for ascertainment, and so continues to produce accurate estimates of h^2^_L_. By contrast, tetrachoric correlation does not allow for ascertainment, and as a result tends to overestimate h^2^_L_ when cases are over-sampled and underestimate h^2^_L_ when cases are under-sampled. For **Figure 4c** we simulate phenotypes where the covariate age explains either 5 or 20% of liability variation. TetraHer is able to accommodate covariates, and therefore continues to produce accurate estimates of h^2^_L_. By contrast, tetrachoric correlation can not include covariates, and as a result tends to underestimate h^2^_L_.

For **Figure 4d**, we simulate phenotypes where 20% of variation in liability is due to common environment (for this we consider the simple case where c[d]=1 for all d, which corresponds to the assumption that all related pairs share a common environment). TetraHer can model this contribution, and therefore continues to produce accurate estimates of h^2^_L_. By contrast, tetrachoric correlation can not model the contribution of common environment, and as a result tends to overestimate h^2^_L_.

The fifth advantage of TetraHer over tetrachoric correlation is that it reports a likelihood, which can be used to perform likelihood ratio tests (i.e., to test whether a trait has significant h^2^_L_ and/or h^2^_C_). **Supplementary Figure 5** indicates that this likelihood is well-calibrated under the null hypothesis.

### Heritability of ICD-10 phenotypes

**Table 1** and **Supplementary Table 1** report estimates of h^2^_L_ for the 229 ICD-10 codes. For these estimates, we assume there is no ascertainment (i.e., that the prevalence in the UK Biobank sample matches the population prevalence), we include covariates (in general, all 23 covariates, however, when analyzing the 37 single-sex codes, we exclude sex) and assume there is no contribution from common environment. 118 of the codes have significant heritability (P<0.05/229 from a likelihood ratio test). These 118 codes span 12 chapters, with 35, 48 and 35 in Levels 1, 2 and 3, respectively (**Supplementary Figure 6**). Instructions on how to construct these phenotypes for UK Biobank individuals are provided on the LDAK website with a summary in **Supplementary Note 5**.

**Table 1:** Significant Level 2 ICD-10 codes. We applied TetraHer to 229 ICD-10 codes, assuming no ascertainment, including 23 covariates, and assuming no contribution from common environment. In total, we identified 118 codes with significant h^2^_L_ (P<0.05/229); this table details the 48 significant Level 2 codes.

| Code | Description | Sex | Prevalence | Estimate of $h^2_L$ | SD |
| --- | --- | --- | --- | --- | --- |
| C44 | Other malignant neoplasms of skin | Both | 0.047 | 0.409 | 0.050 |
| C50 | Malignant neoplasm of breast | Females | 0.063 | 0.413 | 0.069 |
| C61 | Malignant neoplasm of prostate | Males | 0.057 | 0.584 | 0.097 |
| D12 | Benign neoplasm of colon, rectum, anus and anal canal | Both | 0.055 | 0.313 | 0.047 |
| D25 | Leiomyoma of uterus | Females | 0.051 | 0.311 | 0.078 |
| D50 | Iron deficiency anaemia | Both | 0.037 | 0.263 | 0.057 |
| E03 | Other hypothyroidism | Both | 0.054 | 0.447 | 0.045 |
| E11 | Non-insulin-dependent diabetes mellitus | Both | 0.069 | 0.630 | 0.040 |
| E66 | Obesity | Both | 0.062 | 0.415 | 0.043 |
| E78 | Disorders of lipoprotein metabolism and other lipidaemias | Both | 0.138 | 0.351 | 0.031 |
| F17 | Mental and behavioural disorders due to use of tobacco | Both | 0.045 | 0.312 | 0.051 |
| F32 | Depressive episode | Both | 0.054 | 0.265 | 0.048 |
| F41 | Other anxiety disorders | Both | 0.038 | 0.322 | 0.058 |
| G47 | Sleep disorders | Both | 0.021 | 0.360 | 0.089 |
| G56 | Mononeuropathies of upper limb | Both | 0.033 | 0.361 | 0.058 |
| H25 | Senile cataract | Both | 0.053 | 0.283 | 0.051 |
| H26 | Other cataract | Both | 0.070 | 0.324 | 0.048 |
| H40 | Glaucoma | Both | 0.024 | 0.652 | 0.069 |
| I10 | Essential (primary) hypertension | Both | 0.278 | 0.406 | 0.023 |
| I20 | Angina pectoris | Both | 0.059 | 0.328 | 0.048 |
| I25 | Chronic ischaemic heart disease | Both | 0.090 | 0.383 | 0.040 |
| I48 | Atrial fibrillation and flutter | Both | 0.068 | 0.433 | 0.045 |
| I50 | Heart failure | Both | 0.029 | 0.332 | 0.074 |
| I83 | Varicose veins of lower extremities | Both | 0.033 | 0.424 | 0.057 |
| J18 | Pneumonia, organism unspecified | Both | 0.048 | 0.226 | 0.055 |
| J44 | Other chronic obstructive pulmonary disease | Both | 0.039 | 0.445 | 0.060 |
| J45 | Asthma | Both | 0.088 | 0.361 | 0.035 |
| K20 | Oesophagitis | Both | 0.030 | 0.275 | 0.063 |
| K21 | Gastro-oesophageal reflux disease | Both | 0.099 | 0.206 | 0.034 |
| K22 | Other diseases of oesophagus | Both | 0.038 | 0.253 | 0.059 |
| K29 | Gastritis and duodenitis | Both | 0.102 | 0.200 | 0.034 |
| K40 | Inguinal hernia | Males | 0.104 | 0.319 | 0.073 |
| K44 | Diaphragmatic hernia | Both | 0.099 | 0.202 | 0.035 |
| K57 | Diverticular disease of intestine | Both | 0.119 | 0.329 | 0.032 |
| K63 | Other diseases of intestine | Both | 0.063 | 0.268 | 0.044 |
| K80 | Cholelithiasis | Both | 0.050 | 0.248 | 0.049 |
| M16 | Coxarthrosis [arthrosis of hip] | Both | 0.048 | 0.287 | 0.054 |
| M17 | Gonarthrosis [arthrosis of knee] | Both | 0.074 | 0.369 | 0.040 |
| M19 | Other arthrosis | Both | 0.080 | 0.260 | 0.039 |
| M20 | Acquired deformities of fingers and toes | Females | 0.045 | 0.398 | 0.083 |
| M23 | Internal derangement of knee | Both | 0.043 | 0.243 | 0.056 |
| M47 | Spondylosis | Both | 0.035 | 0.251 | 0.062 |
| M48 | Other spondylopathies | Both | 0.020 | 0.344 | 0.087 |
| M51 | Other intervertebral disk disorders | Both | 0.028 | 0.326 | 0.068 |
| M54 | Dorsalgia | Both | 0.057 | 0.236 | 0.047 |
| M79 | Other soft tissue disorders, not elsewhere classified | Both | 0.048 | 0.254 | 0.053 |
| N40 | Hyperplasia of prostate | Males | 0.105 | 0.471 | 0.079 |
| N81 | Female genital prolapse | Females | 0.065 | 0.271 | 0.070 |

We perform three sensitivity analyses. Firstly, we repeat the analysis assuming the population prevalence is twice the sample prevalence, to allow for possible ascertainment due to “healthy volunteer bias”. **Figure 5a** shows that the revised estimates of h^2^_L_ are on average 11% higher than the original ones, while **Figure 5b** shows there is limited change to which codes have significant heritability (three codes change from significant to non-significant, while two change from non-significant to significant). Secondly, we repeat the analysis excluding covariates. **Figure 5c** shows that the revised estimates of h^2^_L_ are similar to the original ones, reflecting that the covariates tend to explain only a small proportion of liability variation (mean 8%, median 6%). Thirdly, we repeat the analysis allowing for the contribution of common environment. **Figure 5d** shows that there is no significant contribution from common environment for any of the 229 codes (the smallest p-value is 0.002, which is not significant after correction for multiple testing).

**Figure 5:**
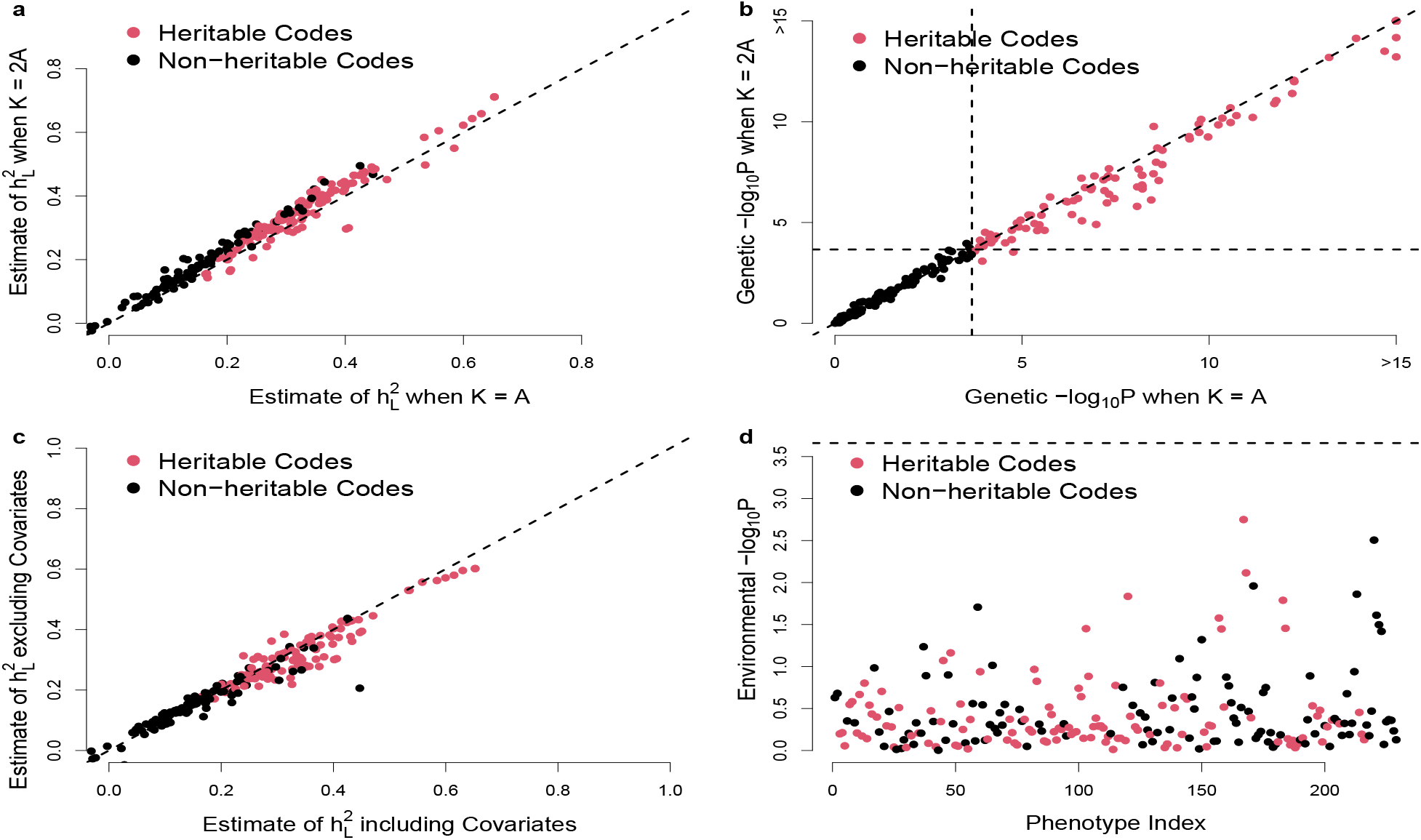
Sensitivity analyses of 229 ICD-10 codes. Our main analysis of the ICD-10 codes assumed no ascertainment, included 23 covariates, and assumed no contribution from common environment. (a) We repeat the analysis assuming the population prevalence is twice the observed prevalence; points compare revised and original estimates of h^2^_L_. (b) Same as (a), except now points compare revised and original -log10 p-values from testing whether h^2^_L_ =0 (note that values above 15 have been truncated). (c) We repeat the analysis excluding covariates; points compare revised and original estimates of h^2^_L_. (d) We repeat the analysis allowing for common environment; points report -log10 p-values from testing whether h^2^_L_ =0. In all panels, red points mark the 118 codes with significant heritability (P<0.05/229) from the original analysis, while horizontal and vertical lines correspond to P=0.05/229.

By way of comparison, we also analyze the quantitative trait height (using QuantHer). As shown in **Supplementary Table 2**, we find a substantial contribution from covariates (in total, they explain 54% of phenotypic variation, primarily driven by sex, and ignoring them reduces the estimate of h^2^_O_ from 0.83 to 0.63). We also find a significant contribution from common environment (the estimate of h^2^_C_ is 0.10, with the likelihood ratio test P<1e-16).

### Relationship between per-SNP heritability and MAF

We hope the 118 significantly heritable ICD-10 codes will be a useful resource for investigating the genetic architecture of human diseases. To provide an example, we use SNP-based heritability analysis to infer the relationship between per-SNP heritability and MAF.^13–15^ Specifically, we use our software SumHer to estimate the power parameter α in the 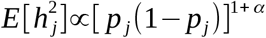 where E[h^2^_j_] is the expected heritability contributed by SNP j, and p_j_ is its MAF.^15,29^ **Figure 6** and **Supplementary Table 3** report estimates of α for the 118 ICD-10 codes. The inverse-variance weighted average estimate across all codes is -0.23 (SD 0.02), and this does not change much if we instead restrict to the 35 Level 1 codes (estimate -0 .19, SD 0.04), the 48 Level 2 codes (estimate -0 .24, SD 0.03) or the 35 Level 3 codes (estimate -0 .26, SD 0.04). While negative α indicates that rarer causal variants tend to have a larger effect size than more common causal variants, consistent with the action of negative selection, we note that the average estimate is higher than that for height (estimate -0.49, SD 0.01). Further, we find only weak evidence that α varies with disease prevalence (weighted least-squares regression slope 0.50, p-value 0.06).

**Figure 6:**
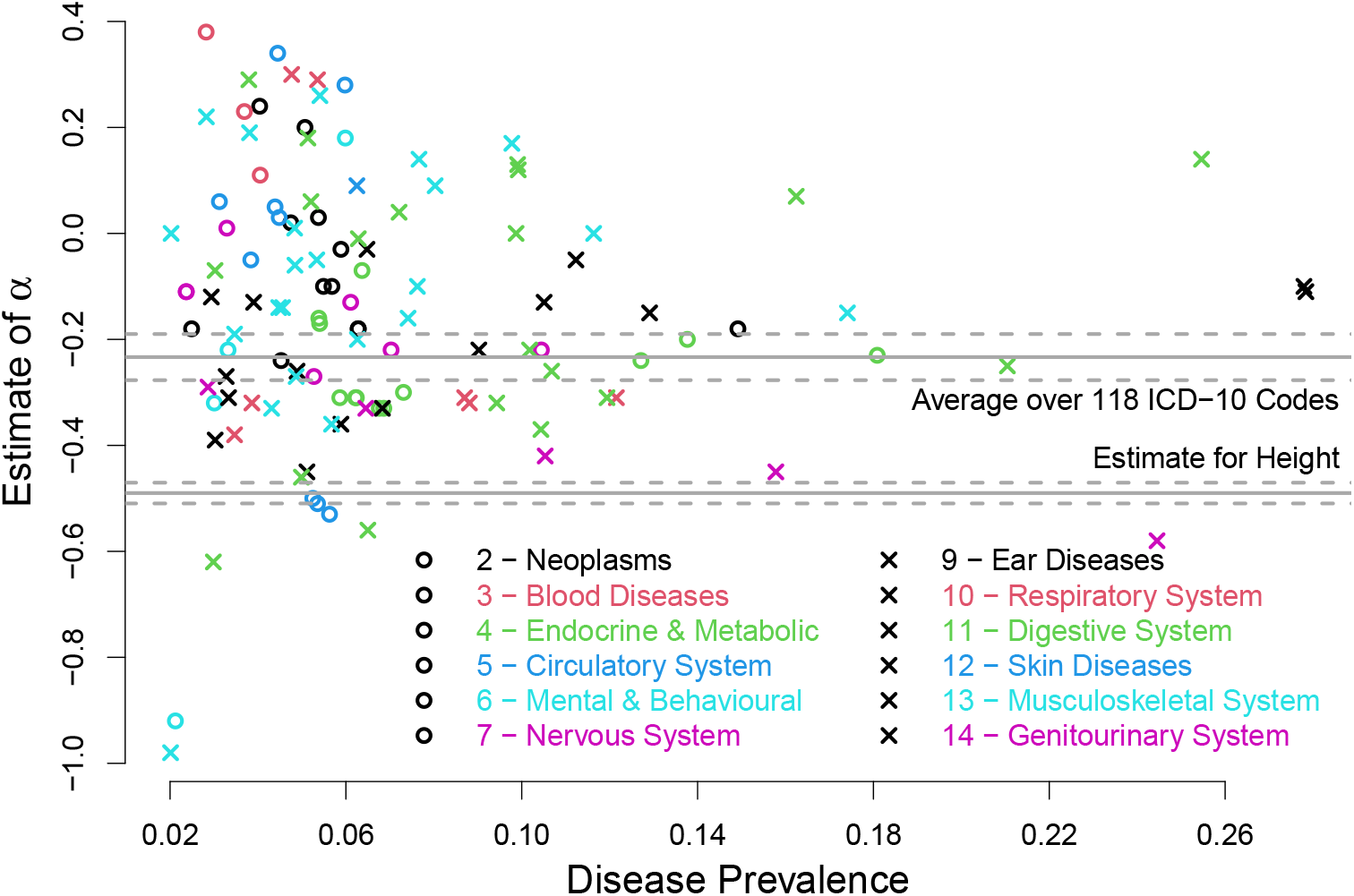
Relationship between per-SNP heritability and MAF. We model the relationship via the power parameter α (see main text). Points report estimates of α for the 118 ICD-10 codes with significant heritability. The disease chapter is indicated by the point shape and color. The top horizontal solid line marks the inverse-variance weighted average across all codes, while the bottom horizontal solid line reports the estimate of α for height (dashed horizontal lines provide the corresponding 95% confidence intervals).

## DISCUSSION

We have developed TetraHer, a method for estimating liability heritability of binary phenotypes which has five key features: it can be applied to complex pedigrees, it allows for ascertainment, it accommodates covariates, it can model the contribution of common environment, and it produces a likelihood.

We first used simulated data to test the validity of TetraHer. We recognize that in our simulations, we had the benefit of knowing the truth, and thus we were able to run TetraHer optimally. In particular, when simulating ascertained phenotypes, we knew the true disease prevalence, while when simulating phenotypes where covariates (common environment) explained liability variation, we knew which covariates to include (the degree of environmental similarity between related pairs). For analyses of real phenotypes, these details are often not available. For example, when analyzing real disease, it can be hard to estimate the population prevalence, while if substantial liability variation is explained by a covariate that is not recorded, TetraHer will be unable to adjust for its contribution.

We additionally used the simulated data to demonstrate the advantage of TetraHer over four existing methods, Pearson’s correlation, PCGC, REML and tetrachoric correlation. We also briefly considered SEM, showing that for the simplest analysis (i.e., where there is only one type of relationship, and ignoring ascertainment, covariates and common environment), estimates from SEM were almost identical to those TetraHer. We recognize that our comparison with SEM was far from comprehensive. This is because there are many implementations of SEM, most of which allow the user to specify a wide variety of models and choose from a range of solvers. In particular, while it is theoretically possible to implement all five features of Tetraher within an SEM framework, doing so would be complicated and require the user to have specialist knowledge. Moreover, we believe that the resulting analyses would be much slower than TetraHer (which always completed within five seconds, even for the most advanced analyses).

We subsequently used TetraHer to identify heritable ICD-10 codes based on UK Biobank data. This analysis demonstrated the advantage of being able to allow for ascertainment, as we could then investigate the potential impact of healthy volunteer bias. Due to the large number of codes analyzed, and the difficulty of finding prevalence estimates for ICD-10 codes, we considered only two scenarios for each code: A=K (i.e., no ascertainment) and K/A=2 (i.e., that the population prevalence was double the sample prevalence). The latter was motivated by a previous study that estimated K/A for four ICD-10 codes with sample prevalence >2% (their estimates were 1.8, 1.8, 2.3 and 2.4), and we consider this a reasonable upper bound for K/A.^30^ While we believe this assumption sufficed in terms of demonstrating estimates are reasonably robust to ascertainment caused by healthy volunteer bias, we recognize that more accurate estimates of h^2^_L_ could be obtained by finding individual estimates of prevalence for each code.

We found that when applying TetraHer to the ICD-10 codes, there was limited advantage including covariates or modeling the contribution of common environment (because neither were estimated to explain a substantial proportion of liability variation). Nonetheless, there are many phenotypes where these two features would be more beneficial (our analysis of height provided one example). Moreover, we expect TetraHer to be advantageous when applied to datasets where inbreeding is common (e.g., for animal and plant datasets), as then there will be a wider spectrum of relatedness values, and therefore a larger benefit being able to use actual relatedness instead of expected relatedness.

We have identified 118 heritable ICD-10 codes, spanning a wide range of disease types, that will be a useful resource for better understanding human diseases. We have provided one example of how this information can be used (to examine the relationship between per-SNP heritability and MAF). However, there are a wide range of other possible applications, such as inferring the number of causal variants, identifying enriched pathways, measuring the performance of prediction models, and estimating genetic correlation between diseases.

## Data Availability

The study used ONLY openly available human data that were originally located at UK Biobank. You can apply for and download the data from https://www.ukbiobank.ac.uk

## ACKNOWLEDGMENTS

D.S. is supported by the Aarhus University Research Foundation (AUFF) and by the Independent Research Fund Denmark (project no. 7025-00094B). D.M.E. is supported by an NHMRC Investigator Grant (application ID 2017942).

## WEB RESOURCES

LDAK, https://www.dougspeed.com

TetraHer documentation, https://www.dougspeed.com/tetraher

Instructions for obtaining the 118 significant ICD10 codes, https://www.dougspeed.com/icd10

