## Supplementary Material for "Estimating disease heritability from complex pedigrees allowing for ascertainment and covariates"

**Supplementary Note 1: Algorithmic details of TetraHer**

**Supplementary Note 2: Existing methods for estimating heritability**

**Supplementary Note 3: Analysis of UK Biobank data**

**Supplementary Note 4: Example scripts for running TetraHer**

**Supplementary Note 5: Instructions for constructing the 118 significant ICD10 diseases**

**Supplementary Figure 1: Comparing TetraHer with polycor**

**Supplementary Figure 2: Comparing polycor with OpenMx**

**Supplementary Figure 3: Comparing existing methods on simulated phenotypes with higher polygenicity**

**Supplementary Figure 4: Comparing existing methods on simulated phenotypes with an alternative effect size distribution**

**Supplementary Figure 5: Calibration of the likelihood ratio test statistics under the null hypothesis**

**Supplementary Figure 6: Estimates of liability heritability for the ICD-10 diseases**

**Supplementary Table 1: Estimates of liability heritability for the 118 significant ICD-10 diseases**

**Supplementary Table 2: Analysis of height**

**Supplementary Table 3: Estimates of the power parameter for the 118 significantly heritable ICD-10 codes**

**Supplementary Note 1: Algorithmic details of TetraHer**

We begin with a reminder of the key notation from the main text. Suppose there are $n$ individuals. Let the length-$n$ vector $Y$ indicate which individuals are affected with a disease, while the length-$n$ vector $L$ contains the corresponding liabilities. Let $A = \sum Y[i]/n$ denote the ascertainment of the disease, and let $K$ denote the prevalence of the disease. Assuming a liability threshold model, it follows that $Y[i] = I(L[i] > T)$, where $T = \Phi^{-1}(K)$ is the (1-$K$)th quantile of a standard normal distribution.[1,2] Lastly, let the $n \times p$ matrix $Z$ contain the covariates, while $R_{i,j}$ and $C_{i,j}$ denote, respectively, estimates of the genetic and environmental similarity between Individuals $i$ and $j$.

Suppose the length-$D$ vectors $S_1$ and $S_2$ contain the indices of $D$ pairs of related individuals. Let $y_1 = Y[S_1]$ and $y_2 = Y[S_2]$ denote the phenotypes for the first and second individuals in each pair, while $l_1 = L[S_1]$ and $l_2 = Y[S_2]$ denote the corresponding liabilities. Further, let the length-$D$ vectors $r$ and $c$ contain the genetic and environmental similarities between pairs. In theory, it is valid to run TetraHer using all possible pairs of individuals (in which case $D =^n C_2$). However, in practice, it is more efficient to restrict to the pairs of individuals with substantial $R_{i,j}$ or $C_{i,j}$ (e.g., with $R_{i,j} \geq 0.05$ or $C_{i,j} \geq 0.05$). This is because pairs with small $R_{i,j}$ and $C_{i,j}$ will have a minimial contribution on the likelihood defined below, so excluding them will have limited impact on the final estimates.

TetraHer assumes the model $L = F + G + C + E$, where the independent, length-$n$ vectors $F$, $G$, $C$ and $E$ denote the contributions of covariates, genetic factors, common environment and environmental noise, respectively. Under this model, the estimate of liability heritability is $h^2_L = Var(G)/(Var(L) - Var(F))$, while $h^2_C = Var(C)/(Var(L) - Var(F))$ is the contribution of common environment. TetraHer is a maximum likelihood method; it first expresses the likelihood of the observed phenotypes as a function of $h^2_L$ and $h^2_C$, then finds $\hat{h^2_L}$ and $\hat{h^2_C}$, the values that maximize the likelihood.

To aid explanation, we first describe the basic version of TetraHer, which assumes there is no ascertainment, and does not allow for either covariates or common environment, then we subsequently explain how each of these three features is incorporated.

**Basic version.** The basic version of TetraHer assumes $L = G + E$ and $A = K$, and only estimates $h^2_L$. To construct a likelihood for the observed phenotypes, it assumes that $(l_1[d], l_2[d])$, the $d$th pair of liabilities, is a draw from a bivariate standard normal distribution with correlation $v[d] = r[d]h^2_L$. It follows that the probabilities of the four possible phenotype pairs are

$$P_{00}[d] = \frac{1}{2\pi\sqrt{1 - v[d]^2}} \int_{-\infty}^{T} \int_{-\infty}^{T} \exp\big(-\frac{a^2 + b^2 - 2v[d]ab}{2(1 - v[d]^2)}\big)\, da\, db,$$

$$P_{10}[d] = \frac{1}{2\pi\sqrt{1 - v[d]^2}} \int_{T}^{\infty} \int_{-\infty}^{T} \exp\big(-\frac{a^2 + b^2 - 2v[d]ab}{2(1 - v[d]^2)}\big)\, da\, db,$$

$$P_{01}[d] = \frac{1}{2\pi\sqrt{1 - v[d]^2}} \int_{-\infty}^{T} \int_{T}^{\infty} \exp\big(-\frac{a^2 + b^2 - 2v[d]ab}{2(1 - v[d]^2)}\big)\, da\, db,$$

$$P_{11}[d] = \frac{1}{2\pi\sqrt{1 - v[d]^2}} \int_{T}^{\infty} \int_{T}^{\infty} \exp\big(-\frac{a^2 + b^2 - 2v[d]ab}{2(1 - v[d]^2)}\big)\, da\, db.$$

Given the above probabilities, we can express the log likelihood for the $d$th phenotype pair as

$$log(P(y_1[d], y_2[d]|v[d])) = I_{00}[d]\, log(P_{00}[d]) + I_{10}[d]\, log(P_{10}[d]) + I_{01}[d]\, log(P_{01}[d]) + I_{11}[d]\, log(P_{11}[d]),$$

where the function $I_{ab}[d]$ indicates whether or not $(y_1[d], y_2[d])$ equals $(a, b)$. TetraHer obtains a joint log likelihood by assuming that pairs of individuals are independent

$$log(P(y_1, y_2|v)) = \sum_d log(P(y_1[d], y_2[d]|v[d])).$$

TetraHer maximizes $log(P(y_1, y_2|v))$ using the Newton-Raphson Method.[3] If $h_L^2[k]$ denotes the estimate of $h_L^2$ at the start of the $k$th iteration, then the revised estimate is

$$h_L^2[k+1] = h_L^2[k] - \frac{dlog(P(y_1, y_2|v))/dh_L^2}{d^2log(P(y_1, y_2|v))/dh_L^{2\,2}},$$

where the numerator and denominator of the fraction are, respectively, the first and second derivatives of the log likelihood evaluated at the current estimate of $h_L^2$. For the first derivative, we have

$$\begin{aligned}
\frac{dlog(P(y_1, y_2|v))}{dh_L^2} &= \sum_d \frac{dlog(P(y_1[d], y_2[d]|v))}{dh_L^2} \\
&= \sum_d \Big( \Big[ I_{00}[d] \frac{dlog(P_{00}[d])}{dv[d]} + I_{10}[d] \frac{dlog(P_{10}[d])}{dv[d]} + I_{01}[d] \frac{dlog(P_{01}[d])}{dv[d]} + I_{11}[d] \frac{dlog(P_{11}[d])}{dv[d]} \Big] \times \frac{dv[d]}{dh_L^2} \Big) \\
&= \sum_d \Big( \Big[ \frac{I_{00}[d]}{P_{00}[d]} \frac{dP_{00}[d]}{dv[d]} + \frac{I_{10}[d]}{P_{10}[d]} \frac{dP_{10}[d]}{dv[d]} + \frac{I_{01}[d]}{P_{01}[d]} \frac{dP_{01}[d]}{dv[d]} + \frac{I_{11}[d]}{P_{11}[d]} \frac{dP_{11}[d]}{dv[d]} \Big] \times r[d] \Big),
\end{aligned}$$

while for the second derivative we have

$$\begin{aligned}
\frac{d^2log(P(y_1, y_2|v))}{dh_L^{2\,2}} = \sum_d \Big( \Big[ &- \frac{I_{00}[d]}{P_{00}[d]^2} (\frac{dP_{00}[d]}{dv[d]})^2 + \frac{I_{00}[d]}{P_{00}[d]} \frac{d^2P_{00}[d]}{dv[d]^2} - \frac{I_{10}[d]}{P_{10}[d]^2} (\frac{dP_{10}[d]}{dv[d]})^2 + \frac{I_{10}[d]}{P_{10}[d]} \frac{d^2P_{10}[d]}{dv[d]^2} \\
&- \frac{I_{01}[d]}{P_{01}[d]^2} (\frac{dP_{01}[d]}{dv[d]})^2 + \frac{I_{01}[d]}{P_{01}[d]} \frac{d^2P_{01}[d]}{dv[d]^2} - \frac{I_{11}[d]}{P_{11}[d]^2} (\frac{dP_{11}[d]}{dv[d]})^2 + \frac{I_{11}[d]}{P_{11}[d]} \frac{d^2P_{11}[d]}{dv[d]^2} \Big] \times r[d]^2 \Big),
\end{aligned}$$

Note that the above expressions for the first and second derivatives both make use of the chain rule (i.e., to obtain the derivative with respect to $h_L^2$, we first differentiate with respect to $v[d]$, then multiply by $dv[d]/dh_L^2 = r[d]$). The derivatives of the probabilities can be computed explicitly. Starting with $P_{00}[d]$, we obtain

$$\begin{aligned}
\frac{dP_{00}[d]}{dv[d]} &= \frac{d}{dv[d]} \left( \frac{1}{2\pi\sqrt{1 - v[d]^2}} \int_{-\infty}^{T} \int_{-\infty}^{T} \exp\big(- \frac{a^2 + b^2 - 2v[d]ab}{2(1 - v[d]^2)}\big)\, da\, db \right) \\
&= \frac{1}{2\pi\sqrt{1 - v[d]^2}} \int_{-\infty}^{T} \int_{-\infty}^{T} \left( \exp\big(- \frac{a^2 + b^2 - 2v[d]ab}{2(1 - v[d]^2)}\big) \times \left[ \frac{v}{1 - v[d]^2} + \frac{ab}{1 - v[d]^2} - \frac{v[d](a^2 + b^2 - 2v[d]ab}{(1 - v[d]^2)^2} \right] \right) da\, db \\
&= \frac{1}{2\pi\sqrt{1 - v[d]^2}} \int_{-\infty}^{T} \int_{-\infty}^{T} \left( \exp\big(- \frac{a^2 + b^2 - 2v[d]ab}{2(1 - v[d]^2)}\big) \times \left[ \frac{v}{1 - v[d]^2} + \frac{(a - v[d]b)(b - v[d]a)}{(1 - v[d]^2)^2} \right] \right) da\, db \\
&= \frac{1}{2\pi\sqrt{1 - v[d]^2}} \exp\big(- \frac{a^2 + b^2 - 2v[d]ab}{2(1 - v[d]^2)}\big) \Big|_{a=T, b=T}. \\
&= \frac{1}{2\pi\sqrt{1 - v[d]^2}} \exp\big(- \frac{T^2(1 - v[d])}{(1 - v[d]^2)}\big).
\end{aligned}$$

Note that the penultimate step (i.e., solving the double integral) involves non-trivial calculus. However, it is (more) straightforward to confirm the step is correct by differentiating the result with respect to $a$ then $b$. Near-identical calculations can be used for the other three probabilities

$$\begin{aligned}
\frac{dP_{10}[d]}{dv[d]} &= -\frac{1}{2\pi\sqrt{1 - v[d]^2}} \exp\big(- \frac{T^2(1 - v[d])}{(1 - v[d]^2)}\big), \\
\frac{dP_{01}[d]}{dv[d]} &= -\frac{1}{2\pi\sqrt{1 - v[d]^2}} \exp\big(- \frac{T^2(1 - v[d])}{(1 - v[d]^2)}\big), \\
\frac{dP_{11}[d]}{dv[d]} &= \frac{1}{2\pi\sqrt{1 - v[d]^2}} \exp\big(- \frac{T^2(1 - v[d])}{(1 - v[d]^2)}\big).
\end{aligned}$$

Note that the differences in sign are a consequence of changing the integration bounds. We can also compute the second derivatives of the probabilities explicitly. Starting with $P_{00}[d]$, we obtain

$$\begin{aligned}\frac{d^2P_{00}[d]}{dv[d]^2} &= \frac{1}{2\pi\sqrt{1-v[d]^2}}\left(\exp\big(-\frac{a^2+b^2-2v[d]ab}{2(1-v[d]^2)}\big)\times\left[\frac{v[d]}{1-v[d]^2}+\frac{(a-v[d]b)(b-v[d]a)}{(1-v[d]^2)^2}\right]\right)\Bigg|_{a=T,b=T}\\ &= \frac{dP_{00}[d]}{dv[d]}\times\left[\frac{v[d]}{1-v[d]^2}+\frac{(a-v[d]b)(b-v[d]a)}{(1-v[d]^2)^2}\right]\Bigg|_{a=T,b=T}\\ &= \frac{dP_{00}[d]}{dv[d]}\times\left[\frac{v[d]}{1-v[d]^2}+\frac{T^2}{(1+v[d])^2}\right].\end{aligned}$$

Near-identical calculations for the other three probabilities result in

$$\begin{aligned}\frac{d^2P_{10}[d]}{dv[d]^2} &= \frac{dP_{10}[d]}{dv[d]}\times\left[\frac{v[d]}{1-v[d]^2}+\frac{T^2}{(1+v[d])^2}\right],\\ \frac{d^2P_{01}[d]}{dv[d]^2} &= \frac{dP_{01}[d]}{dv[d]}\times\left[\frac{v[d]}{1-v[d]^2}+\frac{T^2}{(1+v[d])^2}\right],\\ \frac{d^2P_{11}[d]}{dv[d]^2} &= \frac{dP_{11}[d]}{dv[d]}\times\left[\frac{v[d]}{1-v[d]^2}+\frac{T^2}{(1+v[d])^2}\right].\end{aligned}$$

Having used the Newton-Raphson method to compute $\hat{h^2_L}$, we estimate its variance as

$$Var(\hat{h^2_L}) = \left(\frac{d^2log(P(y_1,y_2|v))}{dh^2_L}\right)^{-1},$$

the inverse of the second derivative of the log likelihood, evaluated at the final estimate. Note that further, technical details of the TetraHer algorithm (e.g., starting estimates) are provided below.

**Allowing for ascertainment.** First we explain how to modify the basic version of TetraHer to allow for ascertainment (i.e., when $A \neq K$). The key modification is that we adjust the probabilities of the four possible phenotype pairs to reflect the differential sampling of cases and controls. Let $q_{case}$ denote the probability that a case randomly picked from the population is included in the study, and let $q_{control}$ denote the corresponding probability for a control. Given that the proportion of cases in the sample is $A$, it follows that

$$q_{control} = \frac{(1-A)K}{A(1-K)}\times q_{case},$$

where the fraction represents the relative chance of including a control compared to a case (it will be less than one if the study is enriched for cases, and vice versa).

We now replace $P_{00}[d]$, $P_{10}[d]$, $P_{01}[d]$ and $P_{11}[d]$ in the equations above with $Q_{00}[d]$, $Q_{10}[d]$, $Q_{01}[d]$ and $Q_{11}[d]$, versions that allow for selection. For example,

$$\begin{aligned}Q_{00}[d] &= \mathbb{P}(y_1[d]=0,y_2[d]=0|\text{Pair } d \text{ in study})\\ &= \frac{\mathbb{P}(\text{Pair } d \text{ in study}|y_1[d]=0,y_2[d]=0)\times\mathbb{P}(y_1[d]=0,y_2[d]=0)}{\mathbb{P}(\text{Pair } d \text{ in study})}\\ &= \frac{{q_{control}}^2\times P_{00}[d]}{\mathbb{P}(\text{Pair } d \text{ in study})}\\ &= \left(\frac{(1-A)K}{A(1-K)}\right)^2 P_{00}[d]\times\frac{{q_{case}}^2}{\mathbb{P}(\text{Pair } d \text{ in study})}.\end{aligned}$$

Note that this makes the assumption that the probability of a pair of controls being included in the study is simply the square of the probability that one control is included. Similar calculations lead to

$$\begin{aligned}Q_{10}[d] &= \frac{(1-A)K}{A(1-K)}P_{10}[d]\times\frac{{q_{case}}^2}{\mathbb{P}(\text{Pair } d \text{ in study})},\\ Q_{01}[d] &= \frac{(1-A)K}{A(1-K)}P_{01}[d]\times\frac{{q_{case}}^2}{\mathbb{P}(\text{Pair } d \text{ in study})},\\ Q_{11}[d] &= P_{11}[d]\times\frac{{q_{case}}^2}{\mathbb{P}(\text{Pair } d \text{ in study})}.\end{aligned}$$

Given that the four probabilities sum to one, we can write

$$
\begin{aligned}
Q_{00}[d] &= \left(\frac{(1-A)K}{A(1-K)}\right)^2 \frac{P_{00}[d]}{s[d]},\\
Q_{10}[d] &= \left(\frac{(1-A)K}{A(1-K)}\right) \frac{P_{10}[d]}{s[d]},\\
Q_{01}[d] &= \left(\frac{(1-A)K}{A(1-K)}\right) \frac{P_{01}[d]}{s[d]},\\
Q_{11}[d] &= \frac{P_{11}[d]}{s[d]},
\end{aligned}
$$

where $s[d] = \left(\frac{(1-A)K}{A(1-K)}\right)^2 P_{00}[d] + \frac{(1-A)K}{A(1-K)} P_{10}[d] + \frac{(1-A)K}{A(1-K)} P_{01}[d] + P_{11}[d]$.

Using these revised probabilities, the log likelihood for the $d$th phenotype pair becomes

$$
log(P(y_1[d], y_2[d]|v[d])) = I_{00}[d]\, log(Q_{00}[d]) + I_{10}[d]\, log(Q_{10}[d]) + I_{01}[d]\, log(Q_{01}[d]) + I_{11}[d]\, log(Q_{11}[d]) - log(s[d]).
$$

The revised first derivative of the joint log likelihood is

$$
\frac{dlog(P(y_1, y_2|v))}{dh_L^2} = \sum_d \left( \left[ \frac{I_{00}[d]}{P_{00}[d]} \frac{dP_{00}[d]}{dv[d]} + \frac{I_{10}[d]}{P_{10}[d]} \frac{dP_{10}[d]}{dv[d]} + \frac{I_{01}[d]}{P_{01}[d]} \frac{dP_{01}[d]}{dv[d]} + \frac{I_{11}[d]}{P_{11}[d]} \frac{dP_{11}[d]}{dv[d]} - \frac{1}{s[d]} \frac{ds[d]}{dv[d]} \right] \times r[d] \right),
$$

while the revised second deriative is

$$
\begin{aligned}
\frac{d^2log(P(y_1, y_2|v))}{{dh_L^2}^2} = \sum_d \Bigg( \Bigg[ &-\frac{I_{00}[d]}{P_{00}[d]^2} (\frac{dP_{00}[d]}{dv[d]})^2 + \frac{I_{00}[d]}{P_{00}[d]} \frac{d^2P_{00}[d]}{dv[d]^2} - \frac{I_{10}[d]}{P_{10}[d]^2} (\frac{dP_{10}[d]}{dv[d]})^2 + \frac{I_{10}[d]}{P_{10}[d]} \frac{d^2P_{10}[d]}{dv[d]^2}\\
&- \frac{I_{01}[d]}{P_{01}[d]^2} (\frac{dP_{01}[d]}{dv[d]})^2 + \frac{I_{01}[d]}{P_{01}[d]} \frac{d^2P_{01}[d]}{dv[d]^2} - \frac{I_{11}[d]}{P_{11}[d]^2} (\frac{dP_{11}[d]}{dv[d]})^2 + \frac{I_{11}[d]}{P_{11}[d]} \frac{d^2P_{11}[d]}{dv[d]^2}\\
&+ \frac{1}{s[d]^2} (\frac{ds[d]}{dv[d]})^2 - \frac{1}{s[d]} \frac{d^2s[d]}{dv[d]^2} \Bigg] \times r[d]^2 \Bigg),
\end{aligned}
$$

where it can be shown that

$$
\frac{ds[d]}{dv[d]} = \left(\frac{(1-A)K}{A(1-K)} - 1\right)^2 \times \frac{dP_{00}[d]}{dv[d]}
$$

and therefore

$$
\frac{d^2s[d]}{dv[d]^2} = \left(\frac{(1-A)K}{A(1-K)} - 1\right)^2 \times \frac{d^2P_{00}[d]}{dv[d]^2}.
$$

**Allowing for common environment.** Next we explain how to modify the basic version of TetraHer to allow for common environment (i.e., $C \neq 0$). The details are almost the same as for the basic version, except that now TetraHer assumes that $(l_1[d], l_2[d])$ is a draw from a bivariate standard normal distribution with correlation $v[d] = r[d]h_L^2 + c[d]h_L^2$, and maximizes the likelihood with respect to both $h_L^2$ and $h_C^2$ using two-dimensional Newton-Raphson iterations. Specifically, if $h_C^2[k]$ denotes the estimate of $h_C^2$ at the start of the $k$th iteration, then the revised estimates of $h_L^2$ and $h_C^2$ are

$$
\begin{pmatrix} h_L^2[k+1] \\ h_C^2[k+1] \end{pmatrix} = \begin{pmatrix} h_L^2[k] \\ h_C^2[k] \end{pmatrix} - \begin{pmatrix} d^2log(P(y_1, y_2|v)/{dh_L^2}^2 & d^2log(P(y_1, y_2|v)/dh_L^2\, dh_C^2 \\ d^2log(P(y_1, y_2|v)/dh_L^2\, dh_C^2 & d^2log(P(y_1, y_2|v)/{dh_C^2}^2 \end{pmatrix}^{-1} \begin{pmatrix} dlog(P(y_1, y_2|v)/dh_L^2 \\ dlog(P(y_1, y_2|v)/dh_C^2 \end{pmatrix}.
$$

The derivatives of the joint log likelihood match those computed in the basic version of TetraHer, except that when differentiating with respect to $h_C^2$, we replace $dv[d]/dh_L^2 = r[d]$ with $dv[d]/dh_C^2 = c[d]$.

**Allowing for covariates.** Lastly we explain how to modify the basic version of TetraHer to allow for covariates (i.e., $F \neq 0$). Let $P[i]$ denote the probability that Individual $i$ is a case, based only on their covariates, and let $K[i]$ denote the probability that a (hypothetical) individual from the population with covariates $Z[i,]$ is a case. The basic version of TetraHer assumes $K[i] = K$, a constant, and thus uses a single liability threshold $T = \Phi^{-1}(K)$. When allowing for covariates, TetraHer copies PCGC[4] and instead uses individual-specific thresholds $T[i] = \Phi^{-1}(\hat{K}[i])$, where $\hat{K}[i]$ is an estimate of $K[i]$. To obtain $\hat{K}[i])$, TetraHer first uses logistic regression to estimate $P[i]$. Specifically, it regresses $Y$ on the covariate matrix $Z$ using the model

$$logit(P[i]) = Z\theta,$$

where the length-$p$ vector $\theta$ contains the coefficients for the covariates. If $\hat{\theta}$ is the estimate of $\theta$ from logistic regression, then the corresponding estimate of $P[i]$ is

$$\hat{P}[i] = \frac{1}{1 - \exp(-Z[i,]\hat{\theta})}.$$

TetraHer subsequently sets

$$\hat{K}[i] = \frac{(1-A)K}{A(1-K)} \Big/ \big(1 + \frac{(1-A)K}{A(1-K)}\hat{P}[i] - \hat{P}[i])\big) \ .$$

The above formula, whose derivation is provided in the PCGC publication, corrects for the fact that when there is ascertainment, $\hat{P}[i]$ is a biased estimate of $K[i]$ (by contrast, when there is no ascertainment, $A = K$, and the above formula reduces to $\hat{K}[i] = \hat{P}[i]$).[4]

The use of individual-specific liability thresholds leads to revised probabilities for the four possible phenotype pairs

$$P_{00}[d] = \frac{1}{2\pi\sqrt{1-v[d]^2}} \int_{-\infty}^{t_1[d]} \int_{-\infty}^{t_2[d]} \exp\big(-\frac{a^2+b^2-2v[d]ab}{2(1-v[d]^2)}\big)\, da\, db$$

$$P_{10}[d] = \frac{1}{2\pi\sqrt{1-v[d]^2}} \int_{t_1[d]}^{-\infty} \int_{-\infty}^{t_2[d]} \exp\big(-\frac{a^2+b^2-2v[d]ab}{2(1-v[d]^2)}\big)\, da\, db$$

$$P_{01}[d] = \frac{1}{2\pi\sqrt{1-v[d]^2}} \int_{-\infty}^{t_1[d]} \int_{t_2[d]}^{-\infty} \exp\big(-\frac{a^2+b^2-2v[d]ab}{2(1-v[d]^2)}\big)\, da\, db$$

$$P_{11}[d] = \frac{1}{2\pi\sqrt{1-v[d]^2}} \int_{t_1[d]}^{-\infty} \int_{t_2[d]}^{-\infty} \exp\big(-\frac{a^2+b^2-2v[d]ab}{2(1-v[d]^2)}\big)\, da\, db,$$

where $t_1 = T[S_1]$ and $t_2 = T[S_2]$ denote the thresholds for the first and second individuals in each pair. Note that the formulae above match those obtained if instead of using individual-specific liability thresholds, we let $f_1 = T - t_1$ and $f_2 = T - t_2$, then assume that $(l_1[d] - f_1[d], l_2[d] - f_2[d])$ has a bivariate standard normal distribution with correlation $v[d]$ (this is the description used in the main text). The first derivatives of the revised phenotype pair probabilities are

$$\frac{dP_{00}[d]}{dv[d]} = \frac{1}{2\pi\sqrt{1-v[d]^2}} \exp\big(-\frac{t_1[d]^2+t_2[d]^2-2v[d]t_1[d]t_2[d]}{2(1-v[d]^2)}\big)$$

$$\frac{dP_{10}[d]}{dv[d]} = -\frac{1}{2\pi\sqrt{1-v[d]^2}} \exp\big(-\frac{t_1[d]^2+t_2[d]^2-2v[d]t_1[d]t_2[d]}{2(1-v[d]^2)}\big)$$

$$\frac{dP_{01}[d]}{dv[d]} = -\frac{1}{2\pi\sqrt{1-v[d]^2}} \exp\big(-\frac{t_1[d]^2+t_2[d]^2-2v[d]t_1[d]t_2[d]}{2(1-v[d]^2)}\big)$$

$$\frac{dP_{11}[d]}{dv[d]} = \frac{1}{2\pi\sqrt{1-v[d]^2}} \exp\big(-\frac{t_1[d]^2+t_2[d]^2-2v[d]t_1[d]t_2[d]}{2(1-v[d]^2)}\big),$$

while the second derivatives are

$$\frac{d^2P_{00}[d]}{dv[d]^2} = \frac{dP_{00}[d]}{dv[d]} \times \left[\frac{v[d]}{1-v[d]^2} + \frac{(t_1[d]-v[d]t_2[d])(t_2[d]-v[d]t_1[d])}{(1-v[d]^2)^2}\right]$$

$$\frac{d^2P_{10}[d]}{dv[d]^2} = \frac{dP_{10}[d]}{dv[d]} \times \left[\frac{v[d]}{1-v[d]^2} + \frac{(t_1[d]-v[d]t_2[d])(t_2[d]-v[d]t_1[d])}{(1-v[d]^2)^2}\right]$$

$$\frac{d^2P_{01}[d]}{dv[d]^2} = \frac{dP_{01}[d]}{dv[d]} \times \left[\frac{v[d]}{1-v[d]^2} + \frac{(t_1[d]-v[d]t_2[d])(t_2[d]-v[d]t_1[d])}{(1-v[d]^2)^2}\right]$$

$$\frac{d^2P_{11}[d]}{dv[d]^2} = \frac{dP_{11}[d]}{dv[d]} \times \left[\frac{v[d]}{1-v[d]^2} + \frac{(t_1[d]-v[d]t_2[d])(t_2[d]-v[d]t_1[d])}{(1-v[d]^2)^2}\right].$$

**QuantHer.** By way of comparison, we also developed and reported results from QuantHer, which estimates heritability for quantitative phenotypes. QuantHer assumes that $(y_1[d], y_2[d])$, the $d$th phenotype pair, has a bivariate standard normal distribution with correlation $v[d] = r[d]h_o^2 + c[d]h_L^2$, which results in the joint log likelihood

$$log(P(y_1, y_2|v)) = \sum_d \frac{1}{2\pi\sqrt{1 - v[d]^2}} \exp\big(-\frac{y_1[d]^2 + y_2[d]^2 - 2v[d]y_1[d]y_2[d]}{2(1 - v[d]^2)}\big).$$

Like TetraHer, QuantHer maximizes $log(P(y_1, y_2|v))$ using the Newton-Raphson Method (except now the primary parameter of interest is $h_O^2$, not $h_L^2$).

**Implementation details.** When using the Newton-Raphson method to maximize the joint log likelihood, our starting estimate of heritability is $h_L^2[0] = 0.5$ (and if allowing for common environment, we set $h_C^2[0] = 0$). By default, the tolerance is 0.001 and the maximum number of iterations is 100 (these can be changed using the options `--tolerance` and `--max-iters`, but in practice, we have not performed an analysis where convergence is an issue). By default, TetraHer permits negative estimates of $h_L^2$ and $h_C^2$. However, the user can constrain estimates to be non-negative by adding `--constrain YES` (note that we do not recommend this restriction, because forcing estimates to be non-negative can result in upwards bias when the true values are close to zero).[5,6]

Let $n' \leq n$ denote the number of individuals that that feature in at least one of the $D$ related pairs. For our analyses of UK Biobank data $n' = n$, reflecting that we selected the $n$ individuals based on relatedness (i.e., we constructed the sample by identifying all individuals who had a close relative also in UK Biobank). However, it is possible to run TetraHer with $n' < n$. In this case, TetraHer will use all $n$ individuals when estimating the contributions of covariates, but when maximizing the joint log likelihood, it will (directly) use only information from the $n'$ individuals in a related pair (note that when there are covariates, the remaining $n - n'$ individuals will indirectly influence the likelihood, because they will affect the estimates of $F$).

**Assumptions underlying TetraHer.** In our view, the primary assumption of TetraHer is that it is appropriate to assume a liability threshold model for the disease being studied (if this is not the case, then estimates of $h_L^2$ become largely meaningless). The validity of the liability model is hard to test. However, we believe it is reasonable provided the disease is polygenic, with no single causal variant of very large effect. This is because, when the genetic affect is spread across a reasonable number of loci, then its distribution across individuals will be approximately normal (a consequence of the Central Limit Theory). For the figure below, we generate liability curves assuming different numbers of causal loci. While it is clear that the liability threshold model is invalid when there is only a single casual locus (because the liabilities of individuals are obviously not normal), it appears reasonable once there are, say, at least five causals (none of which has a very large effect, relative to the others).

Should a user of TetraHer wish to examine the validity of the liability threshold model in more detail, we suggest they first perform a genome-wide association study and estimate the largest proportion of phenotypic variation explained by a single SNP (note that the $\chi^2(1)$ test statistic from linear regression is approximately $nr_j^2$, where $n$ is the number of individuals and $r_j^2$ is the proportion of phenotypic variation explained by the SNP being tested); if this analysis detect a genome-wide significant SNP that explains more than 5% of phenotypic variation, this provides evidence that the liability threshold model is unsuitable (or in the least, that the largest effect SNP should be included as a covariate).

Like most software for estimating heritability, TetraHer assumes an absence of epistasis (i.e., that the contributions of covariates, genetic factors, common environment and environmental noise are independent and that each affects the liability in an additive fashion). Furthermore, when computing the joint log likelihood, TetraHer assumes that the pairs of related individuals are independent. We recognise this assumption is often invalid (e.g., it is automatically false when some individuals feature in more than related pair). We believe that violation of this assumption should not result in biased estimates, but it may affect the corresponding estimates of variance. To explain why, suppose we duplicated all individuals, then ran TetraHer with $2D$ pairs. The resulting estimate of $h_L^2$ would match that from the original analysis (i.e., using $D$ pairs), however, the estimate of its variance would be approximately half that from the original analysis (because

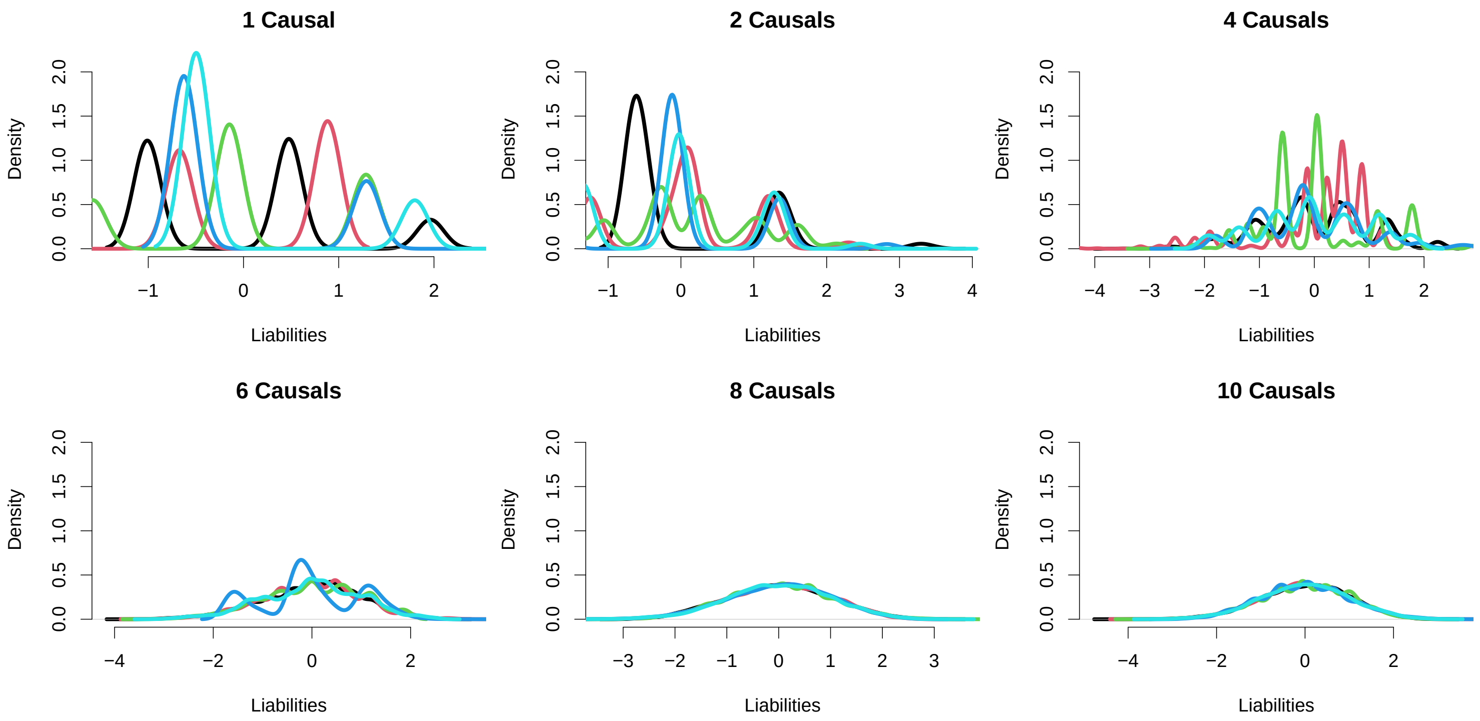


**Normality of Liabilities.** Here we use simple simulations to generate liabilties for $n = 10\,000$ individuals based on $q \in (1, 2, 4, 6, 8, 10)$ causal SNPs. We use the model $L = X\beta$, where the $n \times q$ matrix $X$ contains simulated genotypes for the causal SNPs (for each SNP, genotypes are sampled from a binomial distribution with the minor allele frequency sampled uniformly from $[0.01, 0.5]$), and $\beta$ contains effect sizes sampled from a standard normal distribution. Each panel corresponds to a different value of $q$, and provides density lines for the standardized liabilities across five replicates. When $q = 1$, each set of liabilities takes only three different values (with the heights of the three peaks depending on the minor allele frequency of the causal SNP). As $q$ increases, the liabilities become smoother, such that when $q \geq 6$, they tend to resemble a normal distribution (the exception when $q = 6$, corresponds to a scenario where one of the causal SNPs explains substantially more liability variation than the other five).

TetraHer believes the estimate is supported by twice as much data). To overcome this limitation, TetraHer can instead estimate the variances of estimates via jackknifing (use the option `--num-blocks` to specify the number of partitions). For our analyses of UK Biobank data, we found the jackknife estimates of $Var(\hat{h^2_L})$ were very close to those based on the inverse of the second derivative of the log likelihood, reflecting the near-independence of the pairs of related individuals (e.g., the majority of individuals were included in only one pair).

### Supplementary Note 2: Existing methods for estimating heritability

Here we summarize four existing methods for estimating liability heritability. Note that the first two use a two-step approach, first computing $\hat{h^2_O}$, an estimate of heritability on the observed scale, then converting this to $\hat{h^2_L}$, an estimate of heritability on the liability scale. For this, they use the transformation

$$h^2_L = h^2_O \times \frac{K^2(1-K)^2}{A(1-A)\phi(T)^2},$$

where $\phi(T)$ is the density function of the standard normal distribution (evaluated at $T$, the liability threshold).

**Pearson's correlations.** This obtains $\hat{h^2_O}$ based on the Pearson's correlation between phenotype pairs.[1] It is easiest to use when all pairs have the same relatedness (i.e., $r_d = r_1$ for all $d$), in which case $\hat{h^2_O} = \rho_P / r_1$, where

$$\rho_P = \frac{\sum_d (y_1[d] - \bar{y_1}[d])(y_2[d] - \bar{y_2}[d])}{D-1},$$

the sample correlation across all $D$ pairs. When there are multiple relatedness values, it is in theory possible to group the pairs based on $r_d$, use Pearson's correlations to estimate $h^2_L$ for each group in turn, then combine the resulting estimates into a single estimate (e.g., by computing the inverse-variance weighted mean). However, in practice, this is only feasible when each group is quite large (and so, for example, is not possible when using non-standard estimates of relatedness, as the majority of $r_d$ will be unique). When covariates are provided, we can first regress the phenotypes on these, then compute $\rho_P$ using the residuals.

**REML.** This obtains $\hat{h^2_O}$ using restricted maximum likelihood.[7] First it computes a likelihood by assuming the phenotypes are multivariate normally distributed,

$$Y \sim \mathbb{N}(Z\theta, RVar(G) + IVar(E)),$$

where $R$ is the matrix of pairwise genetic similarities, and $I$ is an $n \times n$ identify matrix. It then obtains a reduced likelihood by integrating across $\theta$. Finally, it computes

$$\hat{h^2_O} = \frac{\hat{Var}(G)}{\hat{Var}(G) + \hat{Var}(E)},$$

where $\hat{Var}(G)$ and $\hat{Var}(E)$ are the values of $Var(G)$ and $Var(E)$, respectively, that maximize the reduced likelihood.

**PCGC.** This first constructs $Y'$, a standardized version of $Y$ such that

$$\mathbb{E}(Y'[i]Y'[j]) = R_{i,j}h^2_L.$$

When there are no covariates, the standardized phenotypes are

$$Y'[i] = \frac{(Y[i] - A)K(1 - K)}{A(1 - A)\phi(T)},$$

while the general form (that allows for covariates), is provided in the PCGC publication.[4] PCGC is a method-of-moment estimator, where $\hat{h^2_L}$ is obtained by regressing $Y'[i]Y'[j]$ on $R_{i,j}$ across all distinct pairs of individuals.

**Tetrachoric correlations.** This obtains $\hat{h^2_O}$ based on the tetrachoric correlation between phenotype pairs.[8] As with Pearson's correlations, it is easiest to use Tetrachoric correlations when all pairs have the same relatedness, in which case $\hat{h^2_L} = \rho_T/r_1$, where $\rho_T$ is the tetrachoric correlation between $y_1$ and $y_2$ (an estimate of the correlation between $l_1$ and $l_2$, the corresponding liabilities). Also as with Pearson's correlations, Tetrachoric correlations can be used when there are multiple relatedness values, by estimating $h^2_L$ for groups with distinct $r_d$, then combining these estimates. We are not aware of any existing methods for estimating Tetrachoric correlations that allow for covariates.

Note that when analyzing quantitative phenotypes, we use Pearson's correlations and REML to estimate $h^2_O$. We omit Tetrachoric correlations, because this can only be applied to binary phenotypes. Then instead of PCGC, we use the analogous method Haseman-Elston Regression (this creates standardized phenotypes, such that $\mathbb{E}(Y'[i]Y'[j]) = R_{i,j}h^2_O$, then obtains $\hat{h^2_O}$ by regressing $Y'[i]Y'[j]$ on $R_{i,j}$).[9]

#### Supplementary Note 3: Analysis of UK Biobank data

We access UK Biobank[10,11] data via Application 21432. In total, UK Biobank provides genotypes and phenotypes for 487 k individuals; our analyses restricted to 56 602 of these, for each of whom there is at least one closely related individual in the data. We selected these individuals as follows. First we filtered the UK Biobank individuals based on ancestry, keeping only the 397 987 who were both recorded and inferred through principal component analysis to be white British. We then identified 121 105 SNPs that are biallelic, have minor allele frequency $> 0.01$, imputation score $> 0.95$ and are in approximate linkage equilibrium (we pruned so no pair within 1 Mb had squared correlation $> 0.05$). Next, we used the software KING (www.kingrelatedness.com) to infer familial relatedness.[12] Specifically, we used the command `king -b ../all.bed --related --degree 2`, where the file `all.bed` contains genotypes (in PLINK format) for the 397 987 individuals and 148 436 SNPs. The main output file, `king.kin0`, details 56 610 individuals who are inferred to have either a sibling, parent or child in the data (the final eight exclusions were due to individuals withdrawing consent since UK Biobank begun). The 56 602 individuals include 32 710 related pairs (142 identical twins, 18 176 full-siblings, 9 398 half-siblings and 4 994 parent-child pairs). For each individual, we have the 23 covariates age, sex, Townsend deprivation index and 20 principal components (ten computed from the UK Biobank data, ten derived from 1000 Genome Project data).

**Simulated phenotypes.** We simulate phenotypes using our software LDAK (www.ldak.org).[6] Suppose the files `data.bed`, `data.fam` and `data.bim` contain genotype data (in PLINK format[13]) for the 56 602 related individuals, each recorded for 121 105 SNPs. To generate the first 600 simulated phenotypes (those underlying Figure 2 in the main text) we use the following script

```
for her in {0.2,0.5,0.8}; do
ldak5.2.linux --make-phenos prev1.her$her.A --her $her --num-causals 1000 --num-phenos 50 \
--bfile data --ignore-weights YES --power -.25 --prevalence 0.01
ldak5.2.linux --make-phenos prev10.her$her.A --her $her --num-causals 1000 --num-phenos 50 \
--bfile data --ignore-weights YES --power -.25 --prevalence 0.1
ldak5.2.linux --make-phenos prev50.her$her.A --her $her --num-causals 1000 --num-phenos 50 \
--bfile data --ignore-weights YES --power -.25 --prevalence 0.5
ldak5.2.linux --make-phenos quant.her$her.A --her $her --num-causals 1000 --num-phenos 50 \
--bfile data --ignore-weights YES --power -.25
done
```

In the above scripts, the first LDAK command generates 50 binary phenotypes. For each phenotype, it first constructs liabilities assuming the model $L = X\beta + e$, where $X$ contains genotypes for 1000 causal SNPs (randomly picked from the 121 105 available), $\beta_j \sim \mathbb{N}(0, [p_j(1-p_j)]^{-0.25}\sigma_g^2)$, where $p_j$ is the minor allele frequency of the $j$th causal SNP, and $e_i \sim \mathbb{N}(0, \sigma_e^2)$. $\sigma_g^2$ and $\sigma_e^2$ are set so that the causal SNPs explain either 20%, 50% or 80% of the variation in $L$. The final phenotypes are $Y = I(L > T)$, where the threshold $T$ assumes the prevalence is 1%. The second and third LDAK commands are the same, except they assume the prevalence is 10% and 50%, respectively. The fourth LDAK command generates quantitative phenotypes, using the model $Y = X\beta + e$ (where $X$, $\beta$ and $e$ are defined the same as above).

The remaining simulations involve binary phenotypes, generated using scripts similar to those above, except that we vary the number of causal SNPs, or change the distribution of $\beta_j$, or filter individuals so as to introduce ascertainment, or allow the covariate age to explain either 5% or 20% of liability variation, or let common environment explain 20% of liability variation.

**Real phenotypes.** For the analysis of real phenotypes, we primarily use the ICD-10 codes provided in field 41270. In total, there are 19 133 ICD-10 codes, which are divided into 22 chapters and four levels: Level 4 codes are a letter followed by three numbers (e.g., A009 denotes unspecified cholera), Level 3 codes are a letter followed by two numbers (e.g., A00 denotes cholera), Level 2 codes are groups of Level 3 codes (e.g., A00-A09 denotes intestinal infectious diseases), while Level 1 codes are chapters (e.g., Chapter 1 denotes infectious and parasitic diseases). Field 41270 specifies which ICD-10 codes have been recorded for each individuals (based on hospital inpatient records). Of the 56 602 related individuals, 6 744 (12%) have no ICD-10 codes recorded, while the remainder have between 3 and 372 recorded (mean 40, median 29). Note that when an individual is assigned an ICD-10 code, they are automatically assigned the corresponding parent and grandparent codes in a hierarchical fashion. For example, if someone is recorded as having the Level 4 code A009, they will also be recorded as having the Level 3 code A00, and the Level 2 code A00-A09, and the Level 1 code Chapter 1.

We restrict to the 229 codes in Chapters 1-15 with prevalence (computed across 251 905 distantly related individuals) at least 2%, of which 65, 90 and 74 are in Levels 1, 2 and 3, respectively. Note that for 37 of the codes, at least 80% of affected individuals were one sex (for 29 codes, females were predominantly affected, while for 8 codes, males were predominantly affected), so for these we exclude individuals of the less-common sex in all analyses. In general, all our analyses of ICD-10 codes assume no ascertainment (i.e., that the prevalence across the related individuals matches the population prevalence), include all 23 covariates, and assume no contribution from common environment (the exception is when we explicitly test the impact of changing one of these three features).

In addition to the ICD-10 codes, we also analyze the quantitative phenotype height (obtained from Field 50).

**Supplementary Note 4: Example scripts for running TetraHer**

Here we summarize the scripts used to run TetraHer. For more detailed instructions, please visit www.dougspeed.com/tetraher. To run the following scripts, you must first download the LDAK executable (visit www.dougspeed.com/downloads). Note that these scripts assume the LDAK executable is called `ldak.out`; you should replace this with either `ldak5.2.linux` or `ldak5.2.mac`, depending on which version of LDAK you have downloaded. You must also download the Test Datasets (visit dougspeed.com/test-datasets). These include four files: the files `disease.relatives` and `disease.enviro` provide details of related individuals; the file `disease.pheno` provides a binary phenotype; while the file `disease.covar` provides covariates. The phenotypes are from a simulated disease with liability heritability 0.5 and prevalence 0.1.

The simplest TetraHer analysis uses the command

```
./ldak.out --family-binary tetraher1 --relatives disease.relatives --pheno disease.pheno
```

By viewing the output file `tetraher1.mle`, we see that the estimated heritability (on the liability scale) is 0.53 (SD 0.05). The phenotype has been ascertained (this is evident from the fact that 20% of the individuals in `disease.pheno` are cases, which is twice the population prevalence). We can allow for this ascertainment using the command

```
./ldak.out --family-binary tetraher2 --relatives disease.relatives --pheno disease.pheno \
--prevalence 0.1
```

Now the estimate of heritability is 0.45 (SD 0.04). To include the covariates in the analysis, we use the command

```
./ldak.out --family-binary tetraher3 --relatives disease.relatives --pheno disease.pheno \
--prevalence 0.1 --covar disease.covar
```

The revised estimate of heritability is 0.56 (0.05). The increase compared to the previous analysis reflects that age is an important covariate (e.g., we see from the file `tetraher3.mle`, that it is estimated to explain 18% of liability variation). To allow for the contribution of common environment, we use the command

```
./ldak.out --family-binary tetraher4 --relatives disease.enviro --pheno disease.pheno \
--prevalence 0.1 --covar disease.covar
```

Now the estimated heritability is 0.32 (SD 0.18), while the estimated contribution of common environment is 0.13 (SD 0.09).

**Supplementary Note 5: Instructions for constructing the 118 significant ICD10 diseases.**

Here we summarize how to construct the significant ICD10 codes for the UK Biobank individuals. These instructions are also available at visit www.dougspeed.com/icd10.

The following scripts use the file icd10.sig (which can be downloaded from www.dropbox.com/s/12nvd5n3eiu029x/icd10.sig), and the UNIX program ukbconv (which is provided by UK Biobank, and can be downloaded from https://biobank.ndph.ox.ac.uk/showcase/download.cgi They also assume you have applied for, received, and decrypted UK Biobank phenotype data (instructions for doing this are provided at https://biobank.ctsu.ox.ac.uk/ bbdatan/Accessing_UKB_data_v2.3.pdf). In this case, you will have a file called ukb23456.enc_ukb, where 23456 will replaced by your run ID..

Step 1. We converted the phenotype data using ukbconv and the commands

```
ukbconv ukb23456.enc_ukb csv -oukb23456
mv ukb23456.enc_ukb.csv ukb23456.csv
```

These commands took about 20 minutes, and the result is a text file called ukb23456.csv.

Step 2. We constructed a file containing ICD10 codes for all UK Biobank individuals (in our case, this file has 502,487 rows and 214 columns). We did this using awk (a built-in UNIX function) and the following three commands

```
head -n 1 ukb23456.csv | awk -v ff=$ff -v FS="\",\"" '{for(j=1;j<=NF;j++){split($j,a,"-"); \
if(a[1]==41270){print j}}}' > extract
W=`wc -l extract | awk '{print $1}'`
awk -v W=$W -v FS="\",\"" '(NR==FNR){a[$1];next}{printf "%s", $1;for (j in a){if($j=="") \
{$j="NA"};printf " %s", $j};printf "\n";}' extract ukb23456.csv | sed s/"\""/""/g > icd10.raw
```

The first command scans the first row of ukb23456.csv (which contains the field IDs) for columns starting with 41270 (which is the field ID for ICD10 codes). In our case, this found 213 columns, which are saved in a file called extract. The second command counts the number of elements of the file extract, and saves this to the variable W. The third command reads all rows of ukb23456.csv, printing out the first column (which contains individual IDs), plus the columns indexed in the file extract. In total, the three commands took about 20 minutes, and the output file is called icd10.raw. If you look at this file (e.g., using the UNIX command `less -S icd10.raw`), you will see most elements are NA. This is OK, and reflects that most individuals have very few (e.g., 0, 1 or 2) of the ICD10 codes.

Step 4. The ICD10 codes are hierarchical. For example, if someone is recorded as having the Level 4 code A009, they also (automatically) have the Level 3 code A00 (the parent of code A009), and the Level 2 code A00-A09 (the parent of code A00). Therefore, to identify cases for each ICD10-defined disease, we must not only find all individuals with the corresponding code, but also individuals with any of the corresponding child codes. The file icd10.sig details the child codes for each ICD10 disease. You can construct a ICD10 disease phenotype using the following awk command

```
for j in {1..118}; do
name=`awk -v num=$j < icd10.sig '(NR==num){print $1}'`
awk -v num=$j < icd10.sig '(NR==num){for(j=1;j<=NF;j++){print $j}}' > pick
echo $j $name
awk '(NR==FNR){a[$1];next}(FNR>1){phen=0;for(j=2;j<=NF;j++){if($j in a){phen=1}}; \
print $1, $1, phen}' pick icd10.raw > $name.pheno
done
```

For each of the 118 diseases in turn, the first commmand obtains the name of the disease, the second extracts the ICD10 codes (both self and child), while the third generates the phenotype file. In total, the command takes about 20 minutes.

The phenotype files are now ready to use with LDAK.

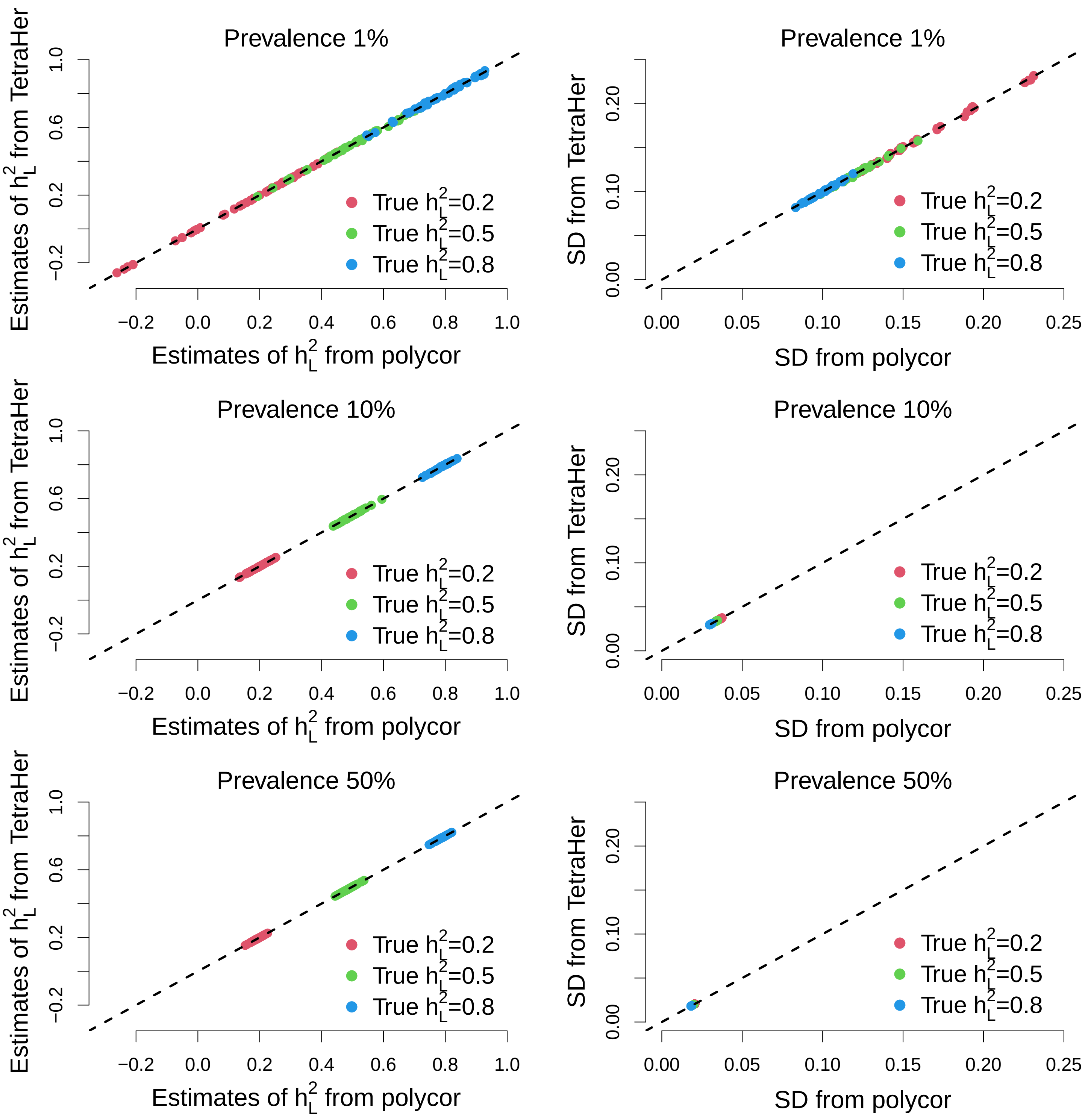


**Supplementary Figure 1: Comparing TetraHer with polycor.** Panels compare estimates of $h_L^2$ from TetraHer ($y$-axis) to those from the R package `polycor` ($x$-axis) for the 450 simulated binary phenotypes underlying Figure 2 in the main text.[14,15] We find near perfect concordance, both for point estimates (Left Column) and the corresponding standard deviations (Right Column).

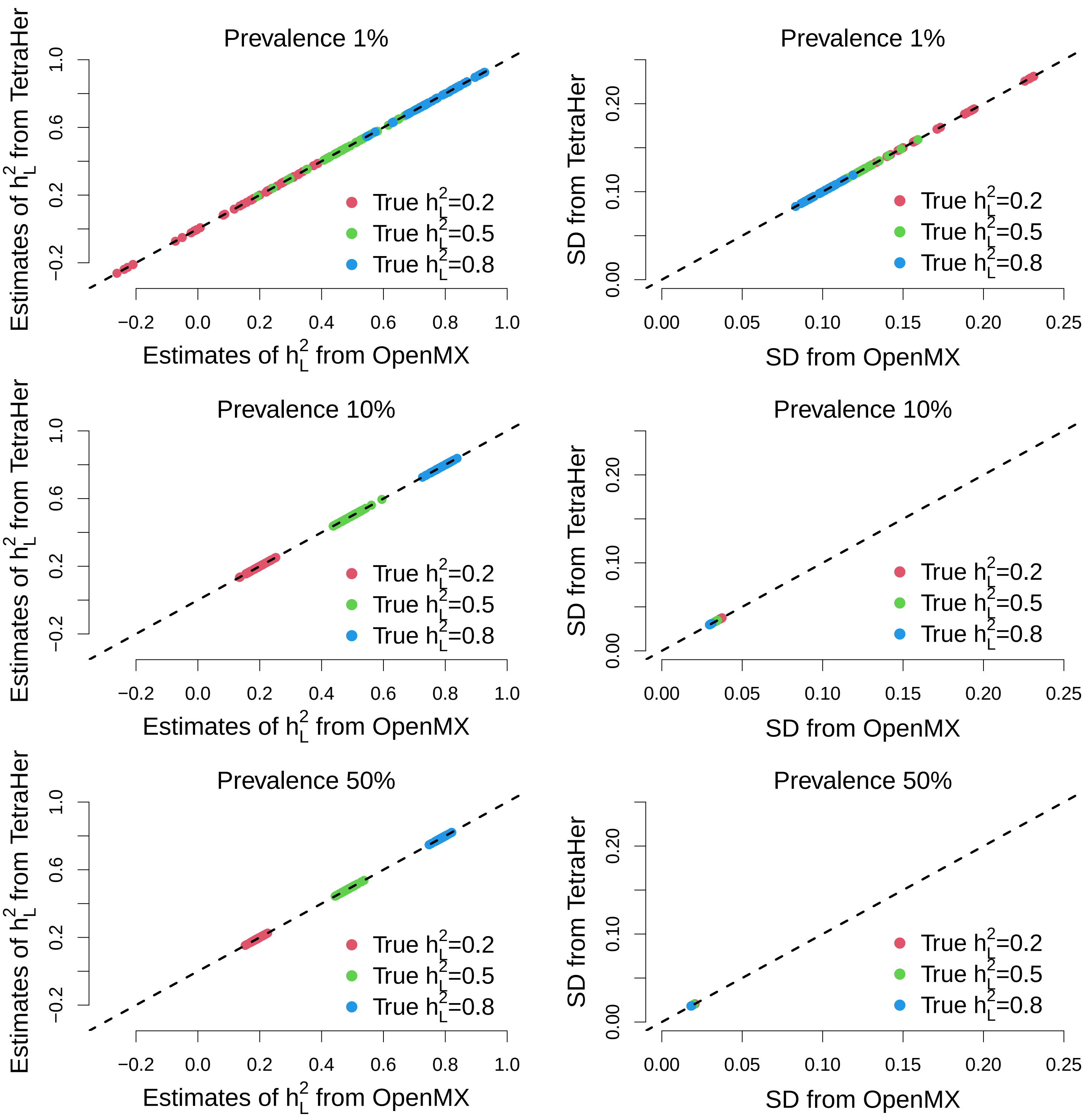


**Supplementary Figure 2: Comparing TetraHer with OpenMx.** Panels compare estimates of $h_L^2$ from TetraHer ($y$-axis) to those from the R package `OpenMx` ($x$-axis) for the 450 simulated binary phenotypes underlying Figure 2 in the main text.[15,16] We find near perfect concordance, both for point estimates (Left Column) and the corresponding standard deviations (Right Column).

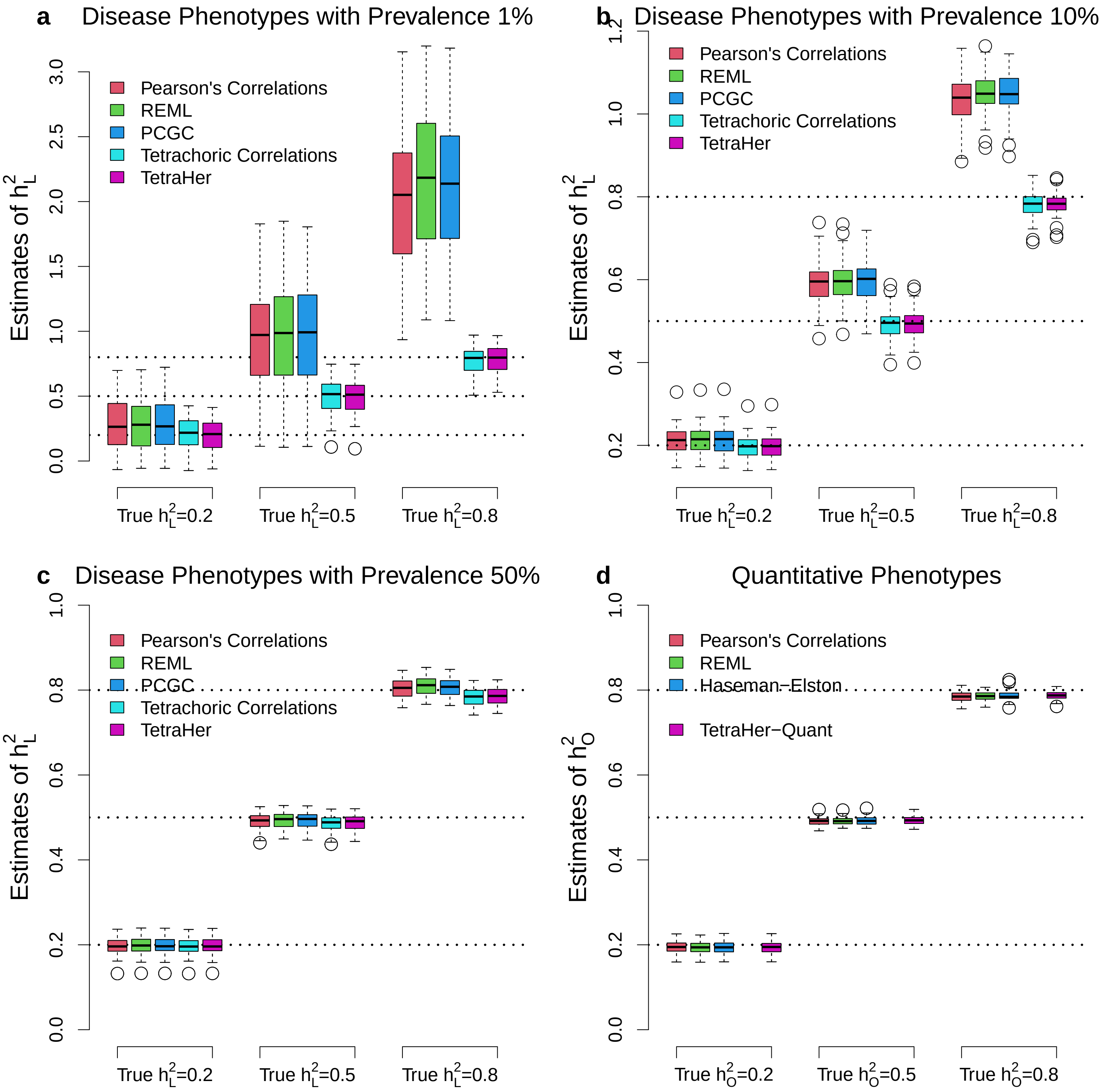


**Supplementary Figure 3: Comparing existing methods on simulated phenotypes with higher polygenicity.** This figure matches Figure 2 in the main text, except that here we increase the number of causal variants from 1000 to 20 000. For full details of the simulation process, see Supplementary Note 3.

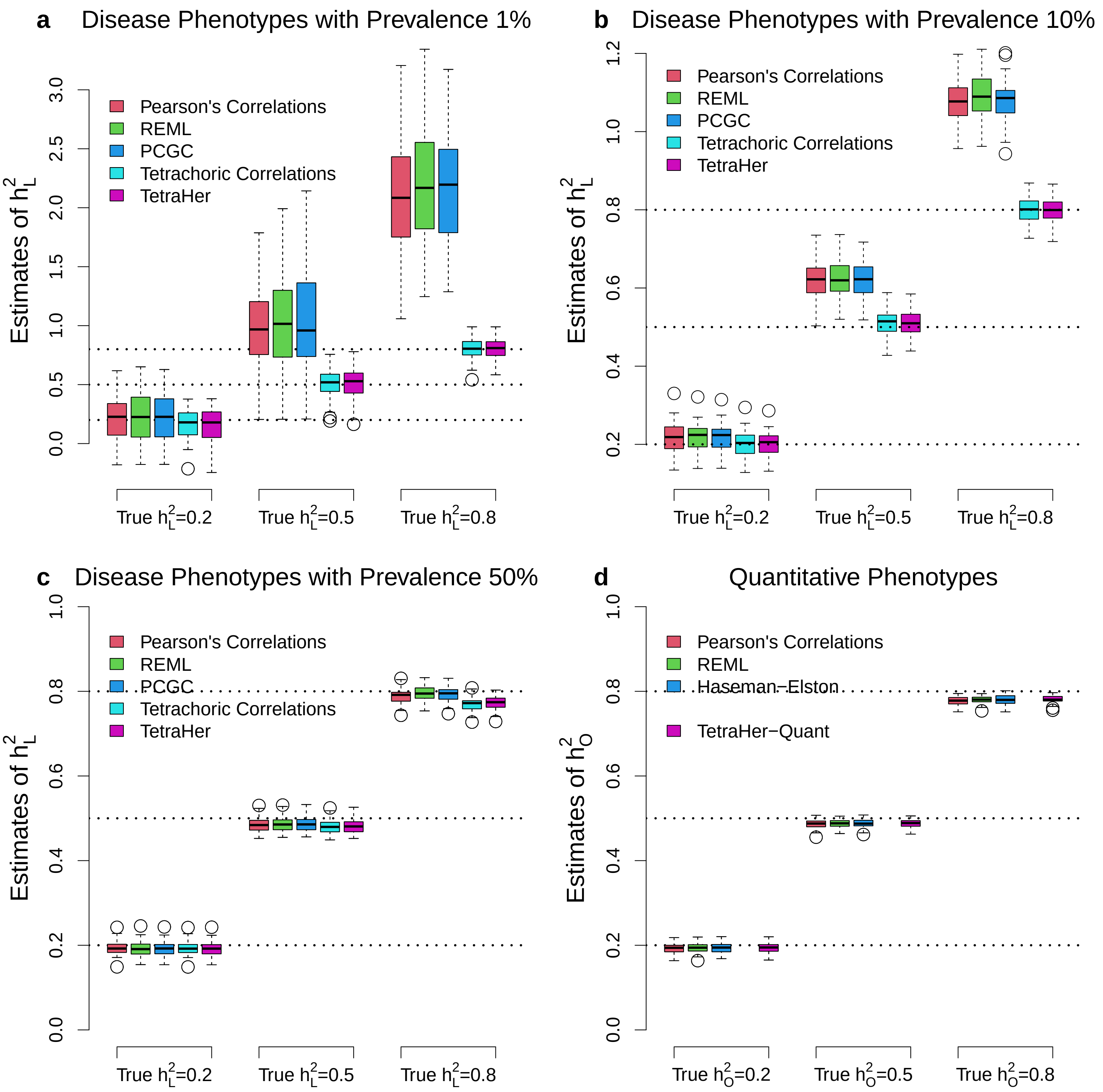


**Supplementary Figure 4: Comparing existing methods on simulated phenotypes with an alternative effect size distribution.** This figure matches Figure 2 in the main text, except that instead of using $\beta_j \sim \mathbb{N}(0, [p_j(1-p_j)]^{-0.25}\sigma_g^2)$, we use $\beta_j \sim \mathbb{N}(0, [p_j(1-p_j)]^{-1}\sigma_g^2)$. For full details of the simulation process, see Supplementary Note 3.

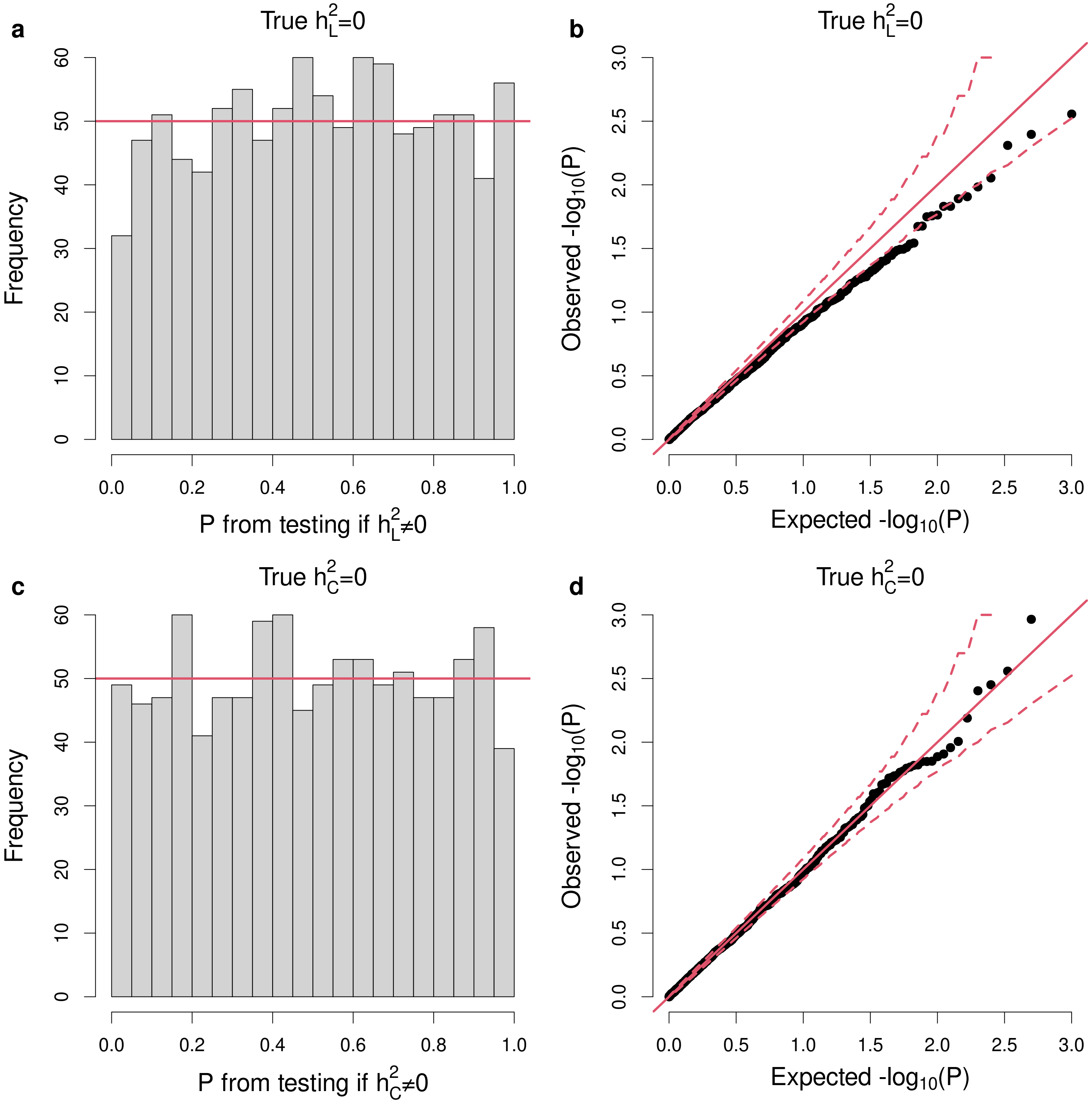


**Supplementary Figure 5: Calibration of the likelihood ratio test statistics under the null hypothesis.** For (a) and (b), we examine the distribution of likelihood ratio test (LRT) statistics when there is no genetic contribution. Specifically, we generate 1000 phenotypes with $h_L^2 = 0$ and $h_C^2 = 0$, analyze these using TetraHer, then generate $p$-values by comparing the LRT statistics to a $\chi^2(1)$ distribution. For (c) and (d), we examine the distribution of LRT statistics when there is no contribution from common environment. For this, we generate 1000 phenotypes with $h_L^2 = 0.2$ and $h_C^2 = 0$, then analyze each phenotype twice, first ignoring common environment, then allowing for it. The LRT statistic is computed as twice the difference in log likelihoods from each pair of analyses (or equivalently, the difference between the pair of LRT statistics). The left plots show the distribution of raw $p$-values, while the right plots show the distribution of ordered -log10 $p$-values. In both scenarios, we find that the observed $p$-values are close the solid red lines, which mark the expected distributions of $p$-values if the LRT statistics are well calibrated (in the right plots, the dashed lines indicate a 95% confidence intervals).

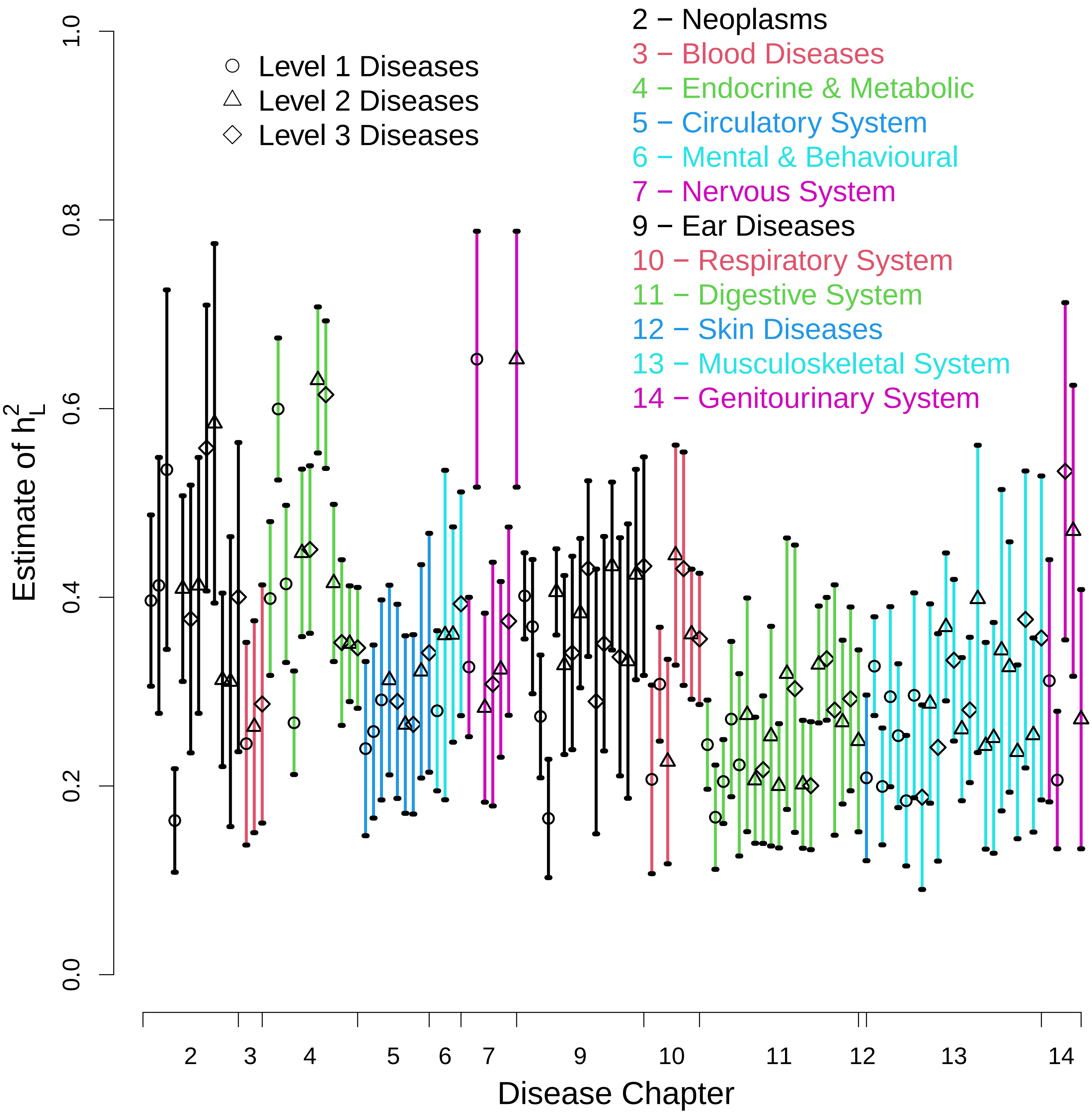


**Supplementary Figure 6: Estimates of liability heritability for the 118 significant ICD-10 diseases.** We applied TetraHer to 229 ICD-10 codes from Chapters 1-15 with prevalence at least 2%. For each analysis, we assumed no ascertainment, included 23 covariates (age, sex, Townsend deprivation index and 20 principal components), and assumed no contribution from common environment. This plot shows the estimates of $h^2_L$ for the 118 codes with significant heritability (P<0.05/229). The shape and colour of points indicate the level of each disease, and to which chapter they belong.

**Supplementary Table 1: Estimates of liability heritability for the ICD-10 diseases.** This table is provided as a spreadsheet in the supplementary tables file. Rows report details for each of the ICD10 codes, as well as estimates of $h^2_L$ and marginal $h^2_L$ (defined as Var(G)/(Var(L)-Var(F)) and Var(G)/Var(L), respectively), and the contribution of covariates (Var(F)/Var(Y)).

| Analysis | Estimate of $h^2_L$ | SD | Estimate of $h^2_C$ | SD | Estimate of $Var(F)/Var(Y)$ | LRT Statistic |
|---|---|---|---|---|---|---|
| **Main Analysis** | 0.828 | 0.015 | 0.102 | 0.009 | 0.539 | 8333 |
| **Ignore Common Environment** | 0.957 | 0.003 | NA | NA | 0.539 | 8187 |
| **Exclude Covariates** | 0.630 | 0.048 | -0.043 | 0.021 | NA | NA |

**Supplementary Table 2: Analysis of height.** Our main analysis of height allows for common environment and includes 23 covariates. Our second analysis includes covariates but ignores common environment, while our third analysis excludes covariates but allows for common environment. When analyzing the ICD-10 codes, there was limited impact when we allowed for common environment, but this is not the case when analyzing height (evidenced by the large estimate of $h^2_C$, and the large drop in the likelihood ratio test (LRT) statistic when ignoring common environment). Similarly, when analyzing the ICD-10 codes, there was limited impact when we included covariates, but this is not the case when analyzing height (evidenced by the large estimate of $Var(F)/Var(Y)$, the proportion of total phenotypic variation explained by covariates).

**Supplementary Table 3: Estimates of the power parameter for the 118 significantly heritable ICD-10 codes.** This table is provided as a spreadsheet in the supplementary tables file. Rows report details for each of the ICD10 codes, as well as estimates of $\alpha$, the parameter which specifies the relationship between $\mathbb{E}[h^2_j]$, the expected heritability contributed by SNP $j$, and $p_j$, its MAF. Formally, these estimates are obtained using SumHer,[17] by assuming the model $\mathbb{E} \propto [p_j(1-p_j)]^{1+\alpha}$, where values of $\alpha$ below (above) -1 indicate that rarer SNPs tend to contribute more (less) heritability than average.
